# Local CCL18 and CCL21 expand lung fibrovascular niches and recruit lymphocytes, leading to tertiary lymphoid structure formation in prolonged COVID-19

**DOI:** 10.1101/2022.03.24.22272768

**Authors:** Ronja Mothes, Anna Pascual-Reguant, Ralf Koehler, Juliane Liebeskind, Alina Liebheit, Sandy Bauherr, Carsten Dittmayer, Michael Laue, Regina von Manitius, Sefer Elezkurtaj, Pawel Durek, Frederik Heinrich, Gitta Anne Heinz, Gabriela Maria Guerra, Benedikt Obermayer, Jenny Meinhardt, Jana Ihlow, Josefine Radke, Frank L. Heppner, Philipp Enghard, Helena Stockmann, Tom Aschman, Julia Schneider, Victor Corman, Leif Erik Sander, Mir-Farzin Mashreghi, Thomas Conrad, Andreas Hocke, Raluca A. Niesner, Helena Radbruch, Anja E. Hauser

**Affiliations:** Department of Neuropathology, Charité–Universitätsmedizin Berlin, corporate member of Freie Universität Berlin, Humboldt-Universität zu Berlin and Berlin Institute of Health, Berlin, Germany; Immune Dynamics, Deutsches Rheuma-Forschungszentrum (DRFZ), a Leibniz Institute, Charitéplatz 1, 10117 Berlin, Germany; Department of Rheumatology and Clinical Immunology, Charité - Universitätsmedizin Berlin, corporate member of Freie Universität Berlin and Humboldt-Universität zu Berlin, 10117 Berlin, Germany; Biophysical Analysis, Deutsches Rheuma-Forschungszentrum (DRFZ), a Leibniz Institute, Berlin, Germany; Centre for Biological Threats and Special Pathogens (ZBS), Robert Koch Institute, Berlin, Germany; Institute of Pathology, Charité–Universitätsmedizin Berlin, corporate member of Freie Universität Berlin, Humboldt-Universität zu Berlin and Berlin Institute of Health, Berlin, Germany; Deutsches Rheuma-Forschungszentrum (DRFZ), a Leibniz Institute, Berlin, Germany; Core Unit Bioinformatics (CUBI), Berlin Institute of Health at Charité-Universitätsmedizin Berlin, Berlin, Germany; Berlin Institute of Health (BIH), Berlin, Germany; German Cancer Consortium (DKTK), Partner Site Berlin, CCCC (Campus Mitte), Berlin, Germany; Cluster of Excellence, NeuroCure, Berlin, Germany; German Center for Neurodegenerative Diseases (DZNE) Berlin, Berlin, Germany; Department of Nephrology and Medical Intensive Care, Charité-Universitätsmedizin Berlin, corporate member of Freie Universität Berlin, Humboldt-Universität zu Berlin, 12203 Berlin, Germany; Institute of Virology, Charité–Universitätsmedizin Berlin, corporate member of Freie Universität Berlin, Humboldt-Universität zu Berlin, and Berlin Institute of Health and German Centre for Infection Research, Berlin, Germany; Department of Infectious Diseases and Respiratory Medicine, Charité, Universitätsmedizin Berlin, Berlin, Germany; German Center for Lung Research (DZL); BIH Center for Regenerative Therapies (BCRT), Charité Universitätsmedizin Berlin, Berlin, Germany; Center for Digital Health, Berlin Institute of Health (BIH) and Charité–Universitätsmedizin Berlin, corporate member of Freie Universität Berlin and Humboldt-Universität zu Berlin, Berlin, Germany; Dynamic and Functional in vivo Imaging, Veterinary Medicine, Freie Universität Berlin, Berlin, Germany

**Keywords:** COVID-19, endothelial dysfunction, fibrovascular niche, heme scavenging, CCL18, CCL21, myofibroblasts, EndMT, lung fibrosis, T cell exhaustion, iBALT

## Abstract

Post-acute lung sequelae of COVID-19 are challenging many survivors across the world, yet the mechanisms behind are poorly understood. Our results delineate an inflammatory cascade of events occurring along disease progression within fibrovascular niches. It is initiated by endothelial dysfunction, followed by heme scavenging of CD163^+^ macrophages and production of CCL18. This chemokine synergizes with local CCL21 upregulation to influence the stromal composition favoring endothelial to mesenchymal transition. The local immune response is further modulated via recruitment of CCR7^+^ T cells into the expanding fibrovascular niche and imprinting an exhausted, T follicular helper–like phenotype in these cells. Eventually, this culminates in the formation of tertiary lymphoid structures, further perpetuating chronic inflammation. Thus, our work presents misdirected immune-stromal interaction mechanisms promoting a self-sustained and non-resolving local immune response that extends beyond active viral infection and leads to profound tissue repurposing and chronic inflammation.

## Introduction

Perivascular niches represent specialized microenvironments in various organs, where immune cells come into contact with stromal cells, in order to exchange information about the state of the tissue and modulate immune responses ^1^. Acute respiratory distress syndrome (ARDS) is the main cause of death in the acute infection phase of COVID-19 ^2^, coursing with severe diffuse alveolar damage (DAD), endothelial disruption and immune cell infiltration in the lungs. These histopathological changes occur as a direct result of SARS-CoV-2 infection ^3^, but also indirectly through a dysregulated immune response ^4^, and lead to a functional breakdown of vasculo-epithelial barriers ^5^. In later disease phases, tissue remodeling mechanisms aimed at repairing the microanatomical lung structure appear compromised in some individuals and lead to tissue scarring and lung fibrosis ^6^, which contribute to chronic respiratory symptoms and fibrotic disease among recovered individuals ^6,7^. Importantly, an aberrant macrophage infiltration, along with increased numbers of fibroblasts in the lung has been proposed as a hallmark of severe COVID-19 disease ^8,9^. In addition, alternatively activated macrophages accumulating in COVID-19 lungs show a pro-fibrotic signature and have been linked to lung fibrosis ^10^, although the exact mechanisms have yet to be clarified. Prolonged changes to the airway immune landscape with increased cytotoxic lymphocytes and B cells, which occur independently of the acute disease severity, have recently been linked to Long-COVID syndrome ^11,12^. Thus, it is of utmost importance to shed light on the recruitment pathways that mediate excessive immune infiltration into the lungs and its implications for disease progression. Whether Long-COVID is the result of a viral reservoir in target organs, or caused by an autoimmune mechanism, or whether both phenomena contribute to the disease is a matter of ongoing discussion. Along with the significance of endothelial dysfunction in the pathomechanisms of COVID-19 disease ^13,14^, with recent reports highlighting post-acute, long-term cardiovascular manifestations ^15^ and chronic pulmonary vascular disease ^7^, the fibrovascular niche deserves special attention to elucidate the role of immune cells for sustained tissue damage.

Here, by combining histopathology, multiplex histology, electron microscopy (EM), three-dimensional light sheet microscopy (LSFM), single-nucleus RNA-sequencing (snSeq) and spatial transcriptomics (ST) we gained insights into the chemokine - chemokine receptor interactions occurring within lung fibrovascular niches during COVID-19. We identify a dysregulated immune-stromal crosstalk that promotes a self-sustained and non-resolving local immune response, extending far beyond active viral infection. Our unique spatial approach identifies endothelial dysfunction as the trigger of aberrant tissue remodeling pathways, leading to an expansion of fibrovascular niches. They emerge as specialized microenvironments hosting T cell aggregates imprinted with exhausted, T follicular helper-like profiles along with bystander activation capabilities, eventually culminating in the formation of induced bronchus-associated lymphoid tissue (iBALT) in prolonged COVID-19 disease.

## Results

### Severe COVID-19 is characterized by progressive lung fibrosis along with an accumulation of immune cells in fibrovascular niches

In order to study the pathophysiological changes that occur in the lung associated to SARS-CoV-2 infection in our cohort, we combined several microscopy techniques and spatial transcriptomics (ST) in consecutive slides of post-mortem lung tissue samples obtained from COVID-19 donors. Lung samples from non-COVID-19-related pneumonia were included as controls. All relevant patient information is shown in Table 1. Hematoxylin/eosin (HE) - staining showed prominent alterations in the tissue composition and microanatomical structure of COVID-19 lungs, characterized by the accumulation of immune cell infiltrates and fibrotic areas that increased with disease duration (Fig. 1A, first row). These phenomena were particularly prominent in late disease stages, culminating in a lack of alveolar spaces. In line with that, the immunofluorescence staining (IF) of adjacent tissue sections acquired by confocal microscopy showed an accumulation of collagen deposits with increasing disease duration, accompanied by an increase of CD45^+^ immune cells (Fig. 1A, second row). While immune cells were scattered throughout the tissue in earlier disease phases, they formed large aggregates at later disease stages. ST analysis of consecutive tissue sections validated the exacerbated collagen deposition at the transcriptional level in samples from later time points (Fig. 1A, third row). We analyzed the same lungs by multi-epitope ligand cartography (MELC) ^16,17^, a multiplex microscopy technique that enabled us to apply a 44-parameter panel for immunofluorescence histology in smaller fields of view (FOV) (Fig. S1A, B). In accordance with the confocal data, we observed an accumulation of collagen deposits and immune cell infiltrates along with disease duration (Fig. 1A, fourth row). Additionally, multiplex microscopy allowed us to allocate the large immune cell aggregates to distinct fibrovascular areas accumulating around smooth muscle actin (αSMA)^+^ structures (Fig. S1C).

**Figure 1.**
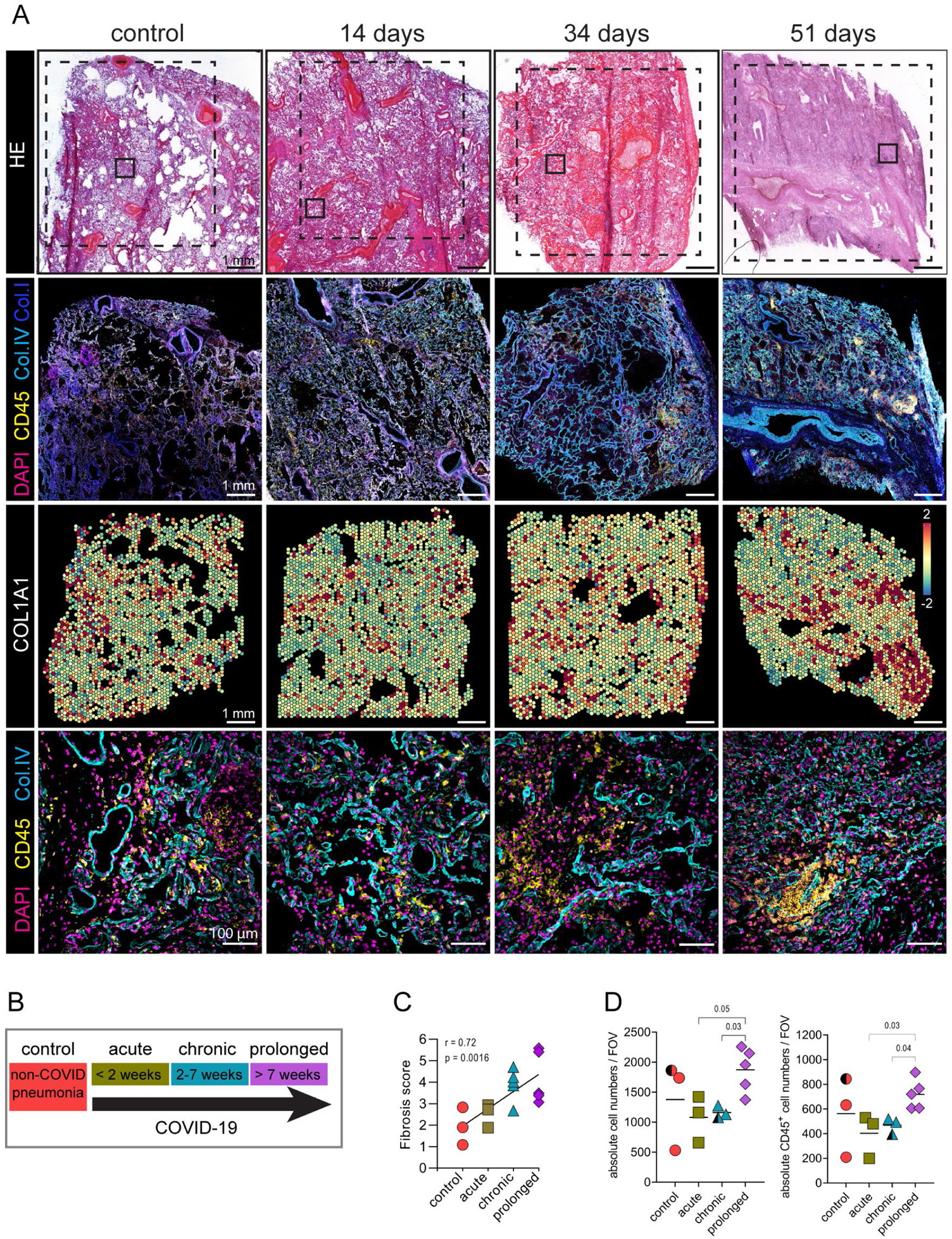
Severe COVID-19 is characterized by progressive lung fibrosis along with an accumulation of immune cells. (A) Consecutive tissue sections of lung samples from non-COVID-related pneumonia donors and COVID-19 donors at different time-points after disease onset were analyzed using spatially resolved techniques. First panel: HE staining shows tissue structure in whole sections with a progressive reduction in alveolar spaces and increased cell infiltration and fibrosis from left to right. Dotted lines represent areas analyzed by confocal microscopy (second panel) and by spatial transcriptomics (ST; third panel). Small squares depict areas analyzed by multiplex microscopy (last panel). Scale bar 1mm. Second panel: corresponding immunofluorescence (IF) images showing CD45 in yellow, DAPI in magenta, Collagen I in cyan and Collagen IV in blue. Third panel: relative expression of *COL1A1* shown as Log2FC and displayed at the spots analyzed with ST. Fourth panel: corresponding IF images showing DAPI in magenta, CD45 in yellow and Collagen IV in cyan. (B) Sample stratification based on disease duration. (C) Dot plot showing the nonparametric Spearman correlation between the stratified samples based on disease duration and fibrosis score evaluated by pathohistological examination of several lung sections from each donor based on Elastica van Gieson staining. See also Table 1. (D) Dot plot depicting the absolute cell numbers (left) and CD45^+^ cell counts (right) per field of view (FOV), which represent 665 × 665 µm, analyzed by multiplex microscopy. Data (mean ± SD) are analyzed by Kruskal-Wallis test with Kruskal-Wallis statistic 6.295 left and 6.707 right. (A, C, D) (n=14). See also Figures S1 and S2.

**Table 1.** Clinical data table.

| Study ID | Age range | Sex | PMI [h] | Disease duration [days] | days at ICU | Invasive ventilation | ECMO | Catecholamines | Corticosteroids | ARDS | Bacterial pneumonia | SARS-CoV2 ante mortem | Mean log10_Sars-CoV-2 GE/10000 diploid cells (LUNG, LN) | Subgenomic RNA (LUNG \| LN) | Nucleocapsid IHC (LUNG) | Prussian Blue (iron) lung stain | Fibrosis score (mean)/of N slides | COVID-19 as part of main cause of death | Technique |
| --- | --- | --- | --- | --- | --- | --- | --- | --- | --- | --- | --- | --- | --- | --- | --- | --- | --- | --- | --- |
| control case 1 | 90-94 | F | 70 | 1 | 1 | yes | no | no | no | no | yes | Negative | N/A \| N/A | N/A \| N/A | 0 | + | 2,83/2 | no | MELC (Lung&LN), ST |
| control case 2 | 55-59 | M | 36 | 55 | 7 | no | no | no | yes | no | yes | Negative | N/A \| N/A | N/A \| N/A | 0 | + | 1,9/1 | no | MELC (Lung), ST |
| control case 3 | 65-69 | M | 40 | 28 | 7 | yes | yes | yes | no | no | yes | Negative | N/A \| N/A | N/A \| N/A | 0 | ++ | 1,08/1 | no | ST |
| control case 4 | 75-79 | M | 50 | N/A | 0 | no | no | no | no | no | yes | N/A | N/A \| N/A | N/A \| N/A | N/A | N/A | N/A | no | snRNAseq |
| acute case 1 | 80-84 | M | 82 | 11 | N/A | no | no | no | no | yes | no | Positive | 6,67 \| 4,85 | yes \| yes | +++ | ++ | 2,93/5 | yes | MELC (Lung), ST, snRNAseq |
| acute case 2 | 90-94 | M | 22 | 12 | 3 | no | no | no | yes | yes | yes | Positive | 5,25 \| 3,11 | yes \| no | ++ | ++ | 1,88/1 | no | MELC (Lung&LN), ST |
| acute case 3 | 75-79 | M | 30 | 13 | 5 | yes | no | yes | yes | no | yes | Positive | 5,89 \| 3,33 | yes \| yes | ++ | NA | 2,73/1 | no | MELC (Lung&LN), ST, snRNAseq, LSFM |
| chronic case 1 | 60-64 | M | 10 | 19 | 13 | yes | no | yes | yes | yes | no | Positive | 2,83 \| 1,33 | no \| no | + | ++ | 2,68/1 | yes | MELC (Lung&LN), ST, snRNAseq |
| chronic case 2 | 65-69 | M | 30 | 34 | 20 | yes | yes | yes | no | yes | yes | Positive | 1,36 \| 2,08 | no \| no | 0 | + | 4,71/4 | yes | MELC (Lung), ST |
| chronic case 3 | 55-59 | F | 38 | 27 | 20 | yes | yes | yes | no | yes | yes | Positive | 1,5 \| 2,02 | no \| no | 0 | 0 | 4,03/5 | yes | MELC (Lung&LN) |
| chronic case 4 | 60-64 | M | 8 | 28 | 0 | no | no | no | no | no | yes | Positive | 2,77 \| 0 | no \| N/A | N/A | NA | 4,23/1 | no | ST |
| chronic case 5 | 65-69 | F | 17 | 34 | 30 | yes | yes | yes | yes | yes | yes | Positive | 0,42 \| 0 | no \| N/A | N/A | N/A | N/A | yes | snRNAseq |
| prolonged case 1 | 70-74 | M | 116 | 51 | 42 | yes | no | yes | no | yes | no | Positive | 1,7 \| 1,37 | no \| no | 0 | +++ | 3,48/5 | yes | MELC (Lung) |
| prolonged case 2 | 75-79 | M | 53 | 51 | 51 | yes | no | yes | yes | yes | no | Positive | 0,58 \| 1,49 | no \| no | 0 | ++ | 3,07/5 | yes | MELC (Lung) |
| prolonged case 3 | 75-79 | M | 84 | 107 | 105 | yes | no | yes | no | no | no | Positive | 0 \| 0 | N/A \| N/A | N/A | NA | 2,62/1 | yes | MELC (Lung&LN), ST, snRNAseq |
| prolonged case 4 | 70-74 | M | 40 | 51 | 48 | yes | yes | yes | no | yes | yes | Positive | 1,9 \| 1,26 | no \| no | 0 | +++ | 5,58/5 | yes | MELC (Lung), ST, LSFM |
| prolonged case 5 | 70-74 | M | 3 | 78 | 72 | yes | yes | yes | no | yes | yes | Positive | 1,31 \| 0 | no \| N/A | 0 | +++ | 5,41/1 | yes | MELC (Lung&LN), ST, LSFM, snRNAseq |

Based on the reported differences correlating with disease duration, we stratified the COVID-19 cases into acute (1 to 15 days of disease duration), chronic (more than 15 days) and prolonged (7 – 15 weeks) (Fig. 1B). Importantly, individuals at the latter phase had resolved infection. Accordingly, lung samples were tested negative or had only very low RNA load by qPCR, targeting the SARS-CoV-2 E gene (Table 1). For active viral replication, subgenomic RNA (sgRNA) was used as a surrogate. We obtained a positive result in all acute cases, but in none of the chronic and prolonged lung tissues analyzed in this study (Table 1). However, both chronic and prolonged cases showed aggravated lung damage.

Next, trained pathologists annotated HE-stained consecutive sections of the ones analyzed by ST in a blinded manner (Fig. S2A). Annotations consisted of intermediate-to-large vessels, airways, alveolar spaces rich in microvasculature as sites of gas exchange, immune infiltrates and fibrotic areas. These annotations served as landmarks for further spatial characterization of the local immune response within distinct functional lung microenvironments. Pathohistological scoring based on HE/Elastica van Gieson (EvG) staining confirmed a high correlation between stratified samples based on disease duration and lung fibrosis (Fig. 1C).

We then pre-processed and segmented the multiplex microscopy immunofluorescence images (see methods) to obtain information of the tissue composition at the single-cell level and be able to quantify cells. While total cell numbers and CD45^+^ cell counts did not differ significantly between controls, acute and chronic COVID-19 groups, there was a significant increase both in cellularity and in absolute numbers of immune cells in the prolonged group (Fig 1D).

Thus, we concluded that severe COVID-19 disease progression in our cohort is characterized by lung fibrosis and that a prominent accumulation of immune cells occurs in prolonged cases, particularly in fibrovascular areas.

### Indirect endothelial damage upon severe COVID-19 induces profound tissue remodeling within fibrovascular niches

Next, we further investigated the endothelial compartment in our cohort. In line with previous studies, we observed drastic lung vasculopathy in severe COVID-19 disease ^5,14^, which presented with a breakdown of the endothelial barrier. CD31 staining was absent in 5 out of 8 FOVs analyzed in the acute lungs but could be detected, although with an aberrant pattern, in later disease phases (Fig. 2A). In line with the previous reports pointing to a failed remodeling of the alveolar epithelium after SARS-CoV-2 infection ^3,5^, we observed a lack in the prototypical monolayer structure in the pancytokeratin (PCK) staining (Fig. 2A). Indicative of progressive lung fibrosis, we also detected an increase in αSMA signal and the reticular fibroblast marker ER-TR7 with disease duration (Fig. 2A). In addition, computational analysis of the multiplexed microscopy image data ^17^, based on dimensionality reduction and unsupervised clustering (Fig. S3A-C), revealed that the absolute numbers of endothelia, epithelia and fibroblasts per FOV were increased, particularly in the prolonged samples (Fig. S3D).

**Figure 2.**
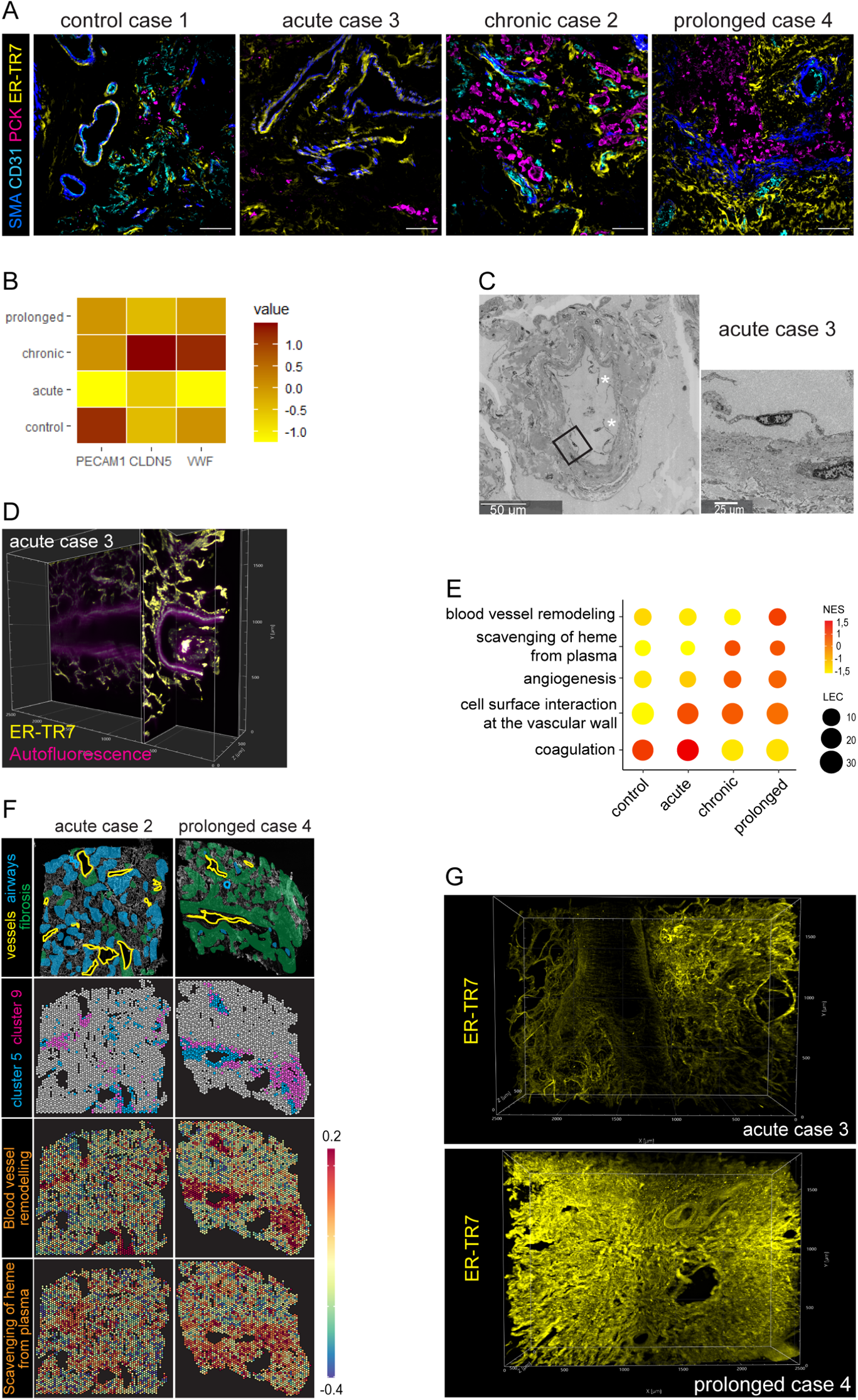
Indirect endothelial damage upon severe COVID-19 induces tissue remodeling mechanisms within fibrovascular niches. (A) Multiplex microscopy immunofluorescence (IF) images depicting smooth muscle actin (αSMA) in blue, CD31 in cyan, pancytokeratin (PCK) in magenta and ER-TR7 in yellow in one representative field of view (FOV) in lung tissue for each disease group. Scale bar 100 µm (n = 32 FOVs). See also Fig. S1 and S3A-D. (B) Heat map display of the average expression level of endothelia-related transcripts in each disease group analyzed by spatial transcriptomics (ST). (C) Ultrastructural image of autopsy lung tissue from one acute donor shows a large vessel with multiple detached endothelial cells within its lumen (white stars). Digitally magnified region of the boxed area shows a detaching endothelial cell. See also Fig. S3E, F. (D) Orthogonal slice of a light sheet fluorescence microscopy (LSFM) acquisition of an acute COVID-19 lung sample stained with ER-TR7 (yellow). Tissue autofluorescence signal (magenta) visualizes a large vessel with a macrothrombus (yellow and magenta) completely clotting it and attached to the vessel wall. See also Video S1. (E) Dot plot depicting the normalized enrichment scores (NES) and leading edge counts (LEC) of transcriptional pathways as analyzed by gene set enrichment analysis (GSEA) of the ST data. The blood vessel remodeling pathway belongs to GOBP, the scavenging of heme from plasma and the cell surface interaction at the vascular wall pathways belong to Reactome and the angiogenesis and coagulation pathways belong to Hallmark. (n = 12 tissue samples). (F) Representative tissue sections of an acute and a prolonged COVID-19 lung samples. First panel: IF image showing collagen I and CD45, both in white, to depict tissue structure and where a blinded and trained pathologist annotated vessels (yellow), airways (blue line), alveolar spaces (blue area) and fibrotic lung areas (green). Second and third panel: color-coded tissue spots analyzed by ST depict the tissue distribution of the NES from the blood vessel remodeling pathway and the scavenging of heme pathway as in (E). (G) LSFM acquisition of an acute and a prolonged COVID-19 lung samples stained with the fibroblast marker ER-TR7 (yellow). See also Video S2 and S3. (B, E, F) (n = 12 tissue samples). (D, G) (n = 4 tissue samples).

Consistent with the loss of CD31 in acutely infected lungs, we detected a downregulation of the endothelial-related transcripts *PECAM1*, *CLDN5* and *VWF* by ST (Fig. 2B) and several vessels with detachment of endothelial cells in the same disease group by electron microscopy (Fig. 2C). The vasculopathy was associated with a prominent coagulopathy, with thrombotic vessels (Fig. 2D, video S1) and coagulation pathways highly abundant in acute lungs (Fig. 2E), as evidenced by gene set enrichment analysis (GSEA) of the ST data. We additionally observed transcriptional pathways of endothelial activation induced upon COVID-19 in the lungs. Heme scavenging and vessel remodeling processes were upregulated in later disease phases (Fig. 2E) in the perivascular areas (Fig. 2F). Further supporting the leakage of plasma from disrupted vessels into the tissue, we detected an increase in Fe^3+^ positive cells (using Prussian blue staining) within COVID-19 lungs along with disease duration (Table 1). Those cells were not restricted to the alveolar space, but also present in perivascular areas. Furthermore, dimensionality reduction and unsupervised clustering analysis of the ST data revealed 12 spot clusters that represented lung microanatomical areas with distinct transcriptional signatures. The clusters were visualized in the UMAP space and the top 25 differentially expressed genes (DEG) were displayed on a heat map (Fig. S4 A-C). Two of these clusters, C5 and C9, represented tissue areas with a stroma-enriched, vascular-associated signature. C5 was characterized by transcripts related to contractility and cytoskeleton rearrangements along with strong expression of *ACTA2,* indicative of intermediate-to-large vessels ^1^. C9, in addition to that, showed the strongest expression of transcripts related to complement activation and endothelial identity. The spatial distribution of these two clusters in all COVID-19 lung samples overlapped with fibrotic landmarks within the perivascular areas, in which heme scavenging and vessel remodeling pathways were particularly enriched (Fig. 2F). Although the fibrotic regions were also most prominent around large vessels, they expanded with disease duration, extending throughout the entire lung parenchyma in the prolonged samples. The acquisition of large image volumes by LSFM further emphasized the profound tissue remodeling occurring during severe COVID-19 disease (Fig. 2G, Video S2, S3). A highly structured, reticular ER-TR7 pattern formed a delicate 3-D scaffold in the acute phase, while it appeared disorganized, dense and compact in the prolonged lungs. Importantly, despite the extensive endothelial dysfunction, we could not find clear signs of pulmonary endothelial infection, neither ultrastructurally (Fig. S3E, F), nor immunohistochemically using antibodies against SARS-CoV-2 nucleocapsid (Fig. S2B).

Hence, we conclude that indirect endothelial dysfunction and leakage in acute COVID-19 lungs lead to heme scavenging along with vessel remodeling within the activated fibrovascular areas, which function as seed-points of fibrosis.

### Dysregulated crosstalk between innate and stromal compartments occurs within fibrovascular niches favoring EndMT

Based on the upregulation of the heme scavenging pathway observed around activated fibrovascular areas, together with the recently demonstrated accumulation of pro-fibrotic macrophages expressing the heme scavenger receptor CD163 in the lungs of severe COVID-19 ^10^, we next interrogated our ST data for the presence of macrophage-derived, pro-fibrotic candidate genes enriched in those activated fibrovascular areas.

One of the clusters that appeared in the ST analysis, C3, showed a macrophage transcriptional signature, characterized by *MARCO*, *CD163*, *HLA-DRA*, *CD68* and *FCGR3A* within the top 25 DEG (Fig. S4A-C). Interestingly, the highest DEG within C3 was *CCL18* (Fig. S4A-C), a chemokine known to be produced by alternatively activated macrophages, inducing fibrosis via crosstalk with fibroblasts in idiopathic pulmonary fibrosis (IPF) ^18^. Single nuclei RNA sequencing (snRNAseq) data obtained from some of the same samples and donors (Table 1) ^19^ demonstrated that CD163^+^ macrophages in COVID-19 are the cellular source of *CCL18* (Fig. S5A). The macrophage population upregulated *CD163* in COVID-19 lungs compared to controls, and showed an enrichment with *CCL18* expression in later disease phases, compared to the acute lungs.

Consistent with the fact that *CD163* and *CCL18* belong to the same ST cluster (C3), the expression of both transcripts co-localized in distinct lung microenvironments that neighbored those areas assigned to C5, C6 and C9 throughout the tissue sections (Fig. S4A-C). These stroma-enriched, vascular-associated clusters were allocated to large vessel landmarks and the fibrotic areas around them (Fig. 3A). While these clusters were transcriptionally distinct from each other, they partly shared a fibrotic signature (Fig. S4C), and could reflect different maturation states of the ongoing fibrosis with a transition from microenvironments characterized by a more endothelial-dominated towards a mesenchymal signature, a process known as endothelial to mesenchymal transition (EndMT) ^20^. Accordingly, C5 showed features of constitutive connective tissue associated with intermediate-to-large vessels ^1^, while C9 also included endothelial-related genes, suggestive of recently generated, capillary-rich fibrous tissue ^21^, and showed a strong expression of *CCL21* (Fig. S4A-C and Fig. 3A)*. CCL21* was hardly detectable in acute COVID-19 samples, but highly upregulated in later disease phases, together with *COL1A1*, *COL3A1* and *COL4A1*, the fibroblast-related transcripts *ACTA2*, *PDGFRA* and *CTHRC1* and the pro-fibrotic mediator *TGF-β* (Fig. S5B). GSEA revealed that the spatial distribution pattern of pathways related to tissue fibrosis mirrored that of *CD163*, *CCL18* and *CCL21* and overlapped with the tissue areas represented by C5, C6 and C9 (Fig. 3 A, B), in contrast to the epithelia-associated clusters C1 and C7 and to C2, which showed an endothelial signature, but lacked contractility-related genes or stromal hallmarks (Fig. 3A and Fig. S4A-C).

**Figure 3.**
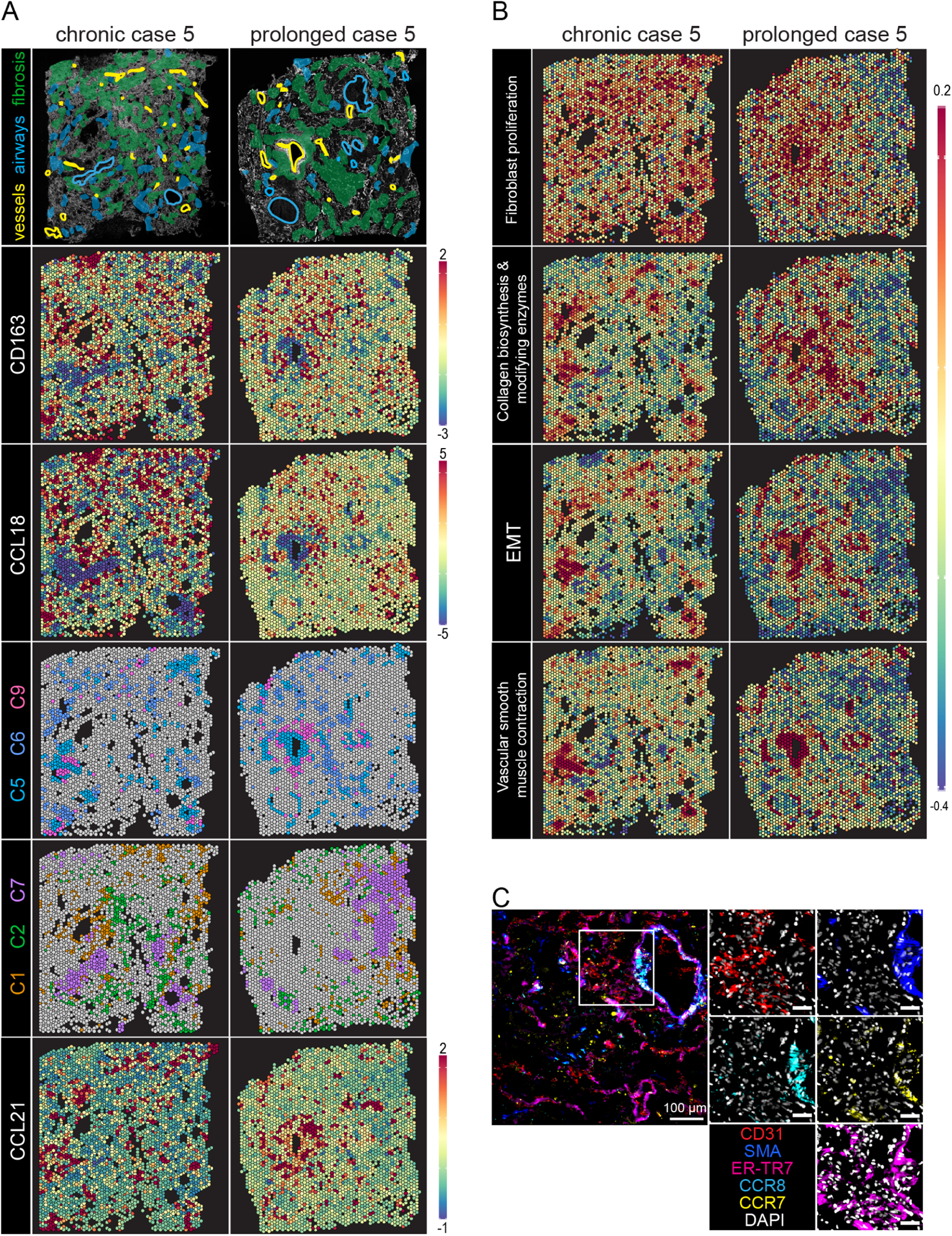
Fibrotic lung areas are enriched in *CD163* and *CCL18* and originate from activated fibrovascular niches with a *CCL21* signature. (A) A representative tissue section from a chronic and a prolonged lung samples. First row: IF image showing collagen I and CD45, both in white, to depict tissue structure and where a blinded and trained pathologist annotated intermediate-to-large vessels (yellow), airways (blue line), alveolar spaces (blue area) and fibrotic lung areas (green) as tissue landmarks. Second and third row: color-coded tissue spots analyzed by ST depict the tissue distribution of the normalized enrichment score (NES) from *CD163* and *CCL18*. Fourth and fifth row: spatial distribution of the color-coded tissue spots depicting relevant ST clusters as shown in Fig. S4. Sixth row: color-coded tissue spots analyzed by ST depict the tissue distribution of the normalized enrichment score (NES) from *CCL21.* (B) Color-coded tissue spots analyzed by ST depict the tissue distribution of the NES from the fibroblast proliferation (GOBP), collagen biosynthesis and modifying enzymes (Reactome), epithelial to mesenchymal transition (Hallmark) and vascular smooth muscle contraction pathways (KEGG). (C) Multiplex microscopy immunofluorescence (IF) overlay depicting αSMA (blue), CCR8 (cyan), CCR7 (yellow), ER-TR7 (magenta) and CD31 (red) in a chronic COVID-19 lung sample. The single channels for each extracellular staining together with DAPI (white) are shown for a region of interest, where scale bar: 40µm (n = 8 FOVs). (A, B) (n = 12 tissue samples).

We next questioned if the activated vasculature was actually able to respond to the CCL18 signal via the cognate receptor CCR8 ^22^, as well as to CCL21 via CCR7. We indeed detected strong CCR8 and CCR7 expression in SMA^+^ and ER-TR7^+^ fibroblasts lining large vessels with disrupted endothelia (Fig. 3C). In addition, several cells of the vascular wall were positive for all three markers, CD31 and SMA and ERTR7, another sign of EndMT ^23–25^. However, while these stromal cells were the main subset expressing the CCR8 receptor within the COVID-19 lung samples analyzed, the CCR7 signal was not restricted to the stromal compartment. Instead, it was also observed in cells with a hematopoietic morphology (Fig. 3C).

Therefore, our results show that activated fibrovascular niches in prolonged COVID-19 lungs host heme scavenging CD163^+^ macrophages, favoring a specialized chemokine milieu including CCL18 and CCL21, which can modulate the local stromal composition and, possibly, the local hematopoietic pool.

### CCR7^+^ T cell aggregates are hosted within activated lung fibrovascular niches of prolonged COVID-19, and imprinted with an unconventional phenotype

We next aimed to further characterize the hematopoietic cells expressing CCR7 within COVID-19 lungs. We detected broad CCR7 expression in large T cells clusters occurring within distinct fibrovascular niches in lung samples from the prolonged group (Fig. 4A, Fig. S6A). Such T cell aggregates were found in 7 out of 12 FOVs of the prolonged lung samples analyzed by multiplex microscopy, while T cells were scattered without forming clusters in the other disease groups. Both CD4^+^ and CD8^+^ T cell counts were significantly enriched in the prolonged lungs (Fig. S3A-C, Fig. 4B and Fig.S6A). LSFM of COVID-19 lungs stained with sytox green and anti-CD3 showed the increase in T cell numbers with disease duration, also in the 3-dimensional space (Video S4 and S5).

**Figure 4.**
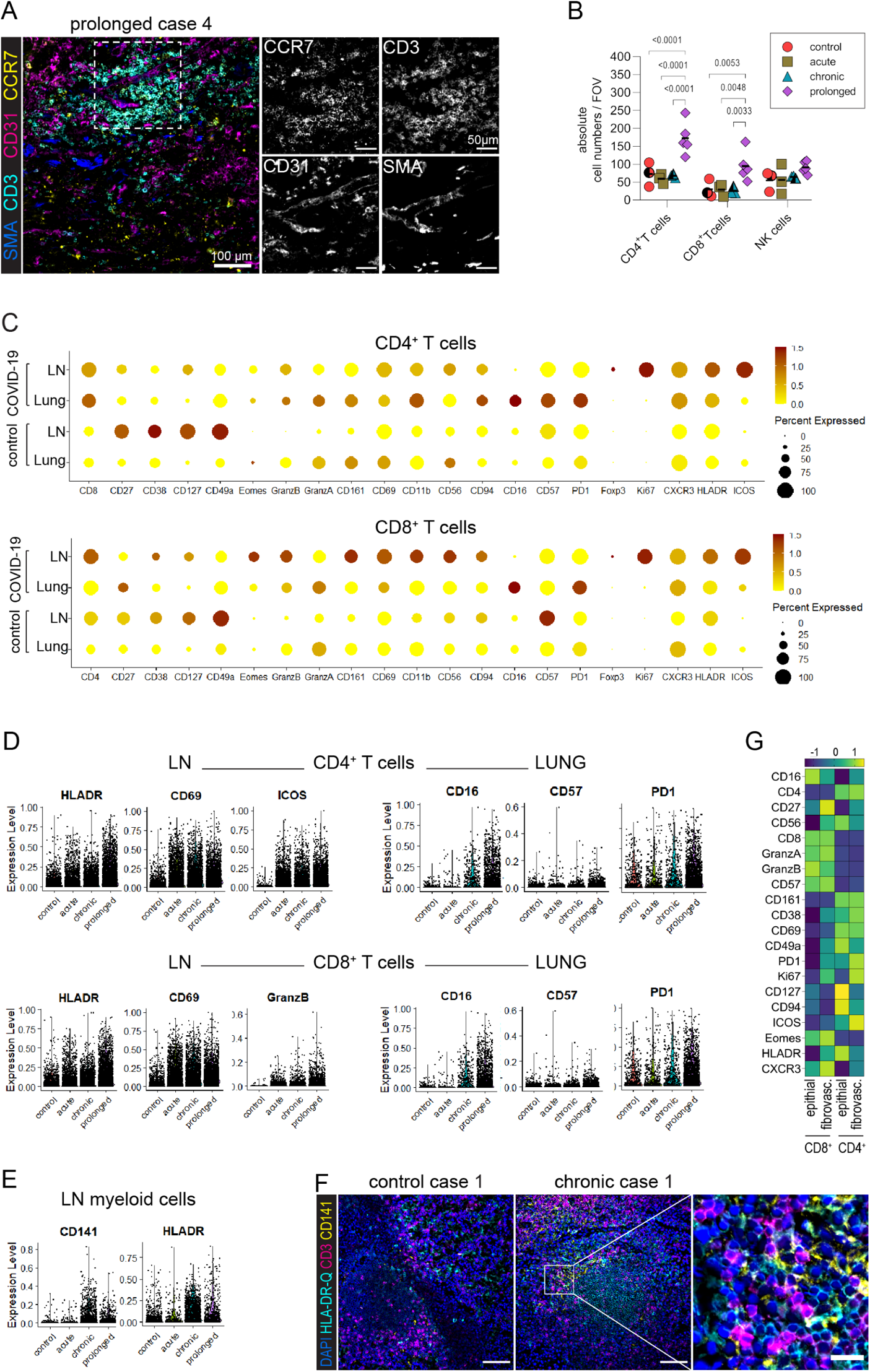
Activated fibrovascular niches of prolonged COVID-19 lungs host CCR7^+^ T cells aggregates and imprint them with an exhausted, T follicular helper-like phenotype. (A) Multiplex microscopy immunofluorescence (IF) image (left) showing CCR7 expression (yellow) in CD3^+^ T cells (cyan) that accumulate in a fibrovascular area marked by αSMA signal (blue) lining CD31^+^ endothelial cells (magenta) in a prolonged COVID-19 sample. The dotted line depicts the region of interest (ROI) for the enlargements. The 4 single channels are shown in white. (n = 8 FOVs). (B) Dot plot representation of the absolute counts per field of view of CD4^+^ and CD8^+^ T cells, as well as NK cells analyzed by multiplex microscopy. Data (mean ± SD) are analyzed by two-way ANOVA with Fisher’s LSD test, where DF = 2 and F = 8.809. See also Fig. S3A-C, Video S4 and S5. (C) Dot plot showing the protein expression profile of CD4+ (upper panel) and CD8+ (lower panel) T cells comparing relative expression (z-score of the mean fluorescence intensity for each marker) in lungs and lymph nodes of COVID-19 donors and controls. Color scale (yellow – dark red) indicates the average expression of the marker and dot size indicates the percent expressed. (n = 41 FOVs). (D) Violin plots show the expression level of HLA-DR-DQ, CD69, ICOS, CD16, CD57 and PD1 in CD4+ (upper panel) and CD8+ (lower panel) T cells in lymph node (left side) and lungs (right side), split by disease group. (E) Violin plots showing the expression level of CD141 and HLA-DR-DQ in the myeloid cells of the lymph nodes per disease group. See also Figure S6 C. (n = 9 FOVs). (F) Immunofluorescence overlays of multiplexed microscopy in lymph nodes depict DAPI (blue), HLA-DR-DQ (cyan), CD3 (magenta) and CD141 (yellow) in a control (left image) and a chronic (middle image) case. The right picture shows the enlargement of the region of interest (white square) from the lymph node tissue of the chronic case (middle image). Scale bars: 100um, 20um in the enlargement image (n = 9 FOVs). (G) Heat map display of the protein expression profile of CD4^+^ and CD8^+^ T cells within the fibrovascular and epithelial tissue niches in the prolonged group analyzed by multiplex microscopy. Data shown as z-score of the mean fluorescence intensity for each marker. See also Figures S1 and S3A-C. (n = 32 FOVs).

Since CCL21 and its receptor CCR7 support T cell migration and homing to lymphoid tissues, but also the recruitment of activated T cells to the lung ^26^, we aimed to compare the local T cell response in the effector site to that of the paired draining lymph nodes (dLNs) (see Table 1 for paired organs analyzed; Fig. S3A-C, Fig. S6A-C and Fig. 4C). CD8 expression within the CD4^+^ T cell population was particularly enriched in COVID-19 lungs, together with several cytotoxicity-related markers. The hallmarks of both CD4^+^ and CD8+ T cells in COVID-19 lungs were broad CD16 and PD1 expression, altogether suggesting an unconventional T cell phenotype. In contrast, both T cell populations in the paired COVID-19 dLNs showed classical markers of activation, such as CD69, HLA-DR-DQ, ICOS and Ki67. On top of that, the expression of Eomes and granzyme B, together with the innate receptors CD16, CD56, CD161 and CD94, was broader and higher in the COVID-19 dLN CD8^+^ T cell subset than in their lung counterparts. The expression level of the antibody-dependent cellular cytotoxicity receptor CD16 and the exhaustion markers CD57 and PD1 in both CD4^+^ and CD8^+^ T cell populations from the lungs increased with disease duration. In parallel, the expression of activation markers in their dLN counterparts increased (Fig. 4D). The upregulation of T cell activation in secondary lymphoid organs in later phases of COVID-19 disease mirrored the increase in CD141 and HLA-DR-DQ expression within the dLN myeloid population (Fig. S6C and Fig. 4E, F). Chronic and prolonged COVID-19 dLNs showed abundant CD141^+^HLADR-DQ^+/−^ dendritic cells (DCs) within the T cell areas, where multiple direct T – DC contacts could be observed (Fig. 4F). Notably, these samples were tested negative or had only very low abundance of *SARS-CoV-2* RNA and no detectable sgRNA, mimicking the qPCR results obtained for the paired lungs (Table 1), thereby indicating that persistent T cell activation within dLNs occurred after infection had been resolved.

Finally, and based on the occurrence of T cell aggregates within fibrovascular niches in the prolonged COVID-19 lungs, we compared the expression profile of T cells in distinct lung niches within those samples (Fig. 4G). We observed the highest expression of ICOS, indicative of a T follicular helper phenotype with increased capability to interact with B cells, and the proliferation marker Ki67, in the CD4^+^ T cell population located within the fibrovascular niche, in comparison to their epithelial niche counterparts. Notably, also CD16 and PD1 expression were upregulated in CD4^+^ T cells within the fibrovascular niche. Co-expression of all these markers was confirmed at the single cell level within those areas (Fig. S6A), highlighting the role of this microenvironment for imprinting an unconventional T cell phenotype.

Altogether, we conclude that while T cell activation persists in the draining lymph nodes in prolonged COVID-19 disease, lung fibrovascular niches, which expand and upregulate CCL21, provide a space where CCR7^+^ T cells accumulate and acquire an unconventional, exhausted T follicular helper-like phenotype with proliferative capacities.

### CCL21^+^ fibrovascular niches aim at the formation of tertiary lymphoid structures in prolonged COVID-19 lungs

Some of the large immune cell aggregates observed in the prolonged lung samples appeared particularly densely packed, expressed high levels of CD45, were associated to collagen^+^ structures in the fibrovascular space and sometimes, directly attached to bronchi / bronchioli, as shown in (Fig. 5A). Immune cells contained in this airway were also expressing CD45, albeit at lower levels than the cells in the fibrovascular space. ST analysis revealed a unique transcriptional fingerprint within the area where the CD45^hi^ cell cluster was located (Fig. 5A, B). In line with our multiplex microscopy results, T cell-specific transcripts such as *CD3D* and *TCF7* accumulated within that area, where also *MKI67* was transcribed and where *CCL21* was also enriched (Fig. 3A, lower row). In addition, *CCL19*, *LTB* and *CXCL13* (Fig. 5B) were particularly elevated in and restricted to that area. These cytokines play a crucial role in the organization of lymphoid structures ^27^. Along that line, the B cell master regulator *PAX5* and *CCR6*, which has been related to B cell activation and memory formation ^28,29^, were exclusively expressed there (Fig. 5B). *IL7R*, *HLA-DRA* transcripts were broader expressed across the tissue section, but they were also found enriched within that region (Fig. 5B), indicating the presence of survival cues and occurrence of antigen presentation. Of note, macrophage-related transcripts, such as *MARCO* and *IL1B were* not expressed within the lymphocyte-enriched area, but rather within the infiltrated airway structures (Fig. S7). On the other hand, the master regulator of plasma cell identity *PRDM1,* together with the immunoglobulin transcripts *IGHG1*, *IGHD*, *IGHA1* and I*GHM* were very prominently expressed in airway-surrounding regions and lower levels of *IGHD*, *IGHA1* and I*GHM* were found within the lymphoid structure (Fig. 5B). While *IGHA1* was robustly detected within all lung samples analyzed, *IGHA2* transcripts were mainly absent, in accordance with published data from blood ^30^ and lung ^31^. Similar large, dense and structured immune cell aggregates, consisting of both T and B cells, could be observed in 2 out of 3 prolonged lungs analyzed by ST. Consistently, we also observed a significant increase in the absolute numbers of B cells / plasma cells in the prolonged COVID-19 group compared to control tissues and earlier disease stages by multiplex microscopy (Fig. 5C).

**Figure 5.**
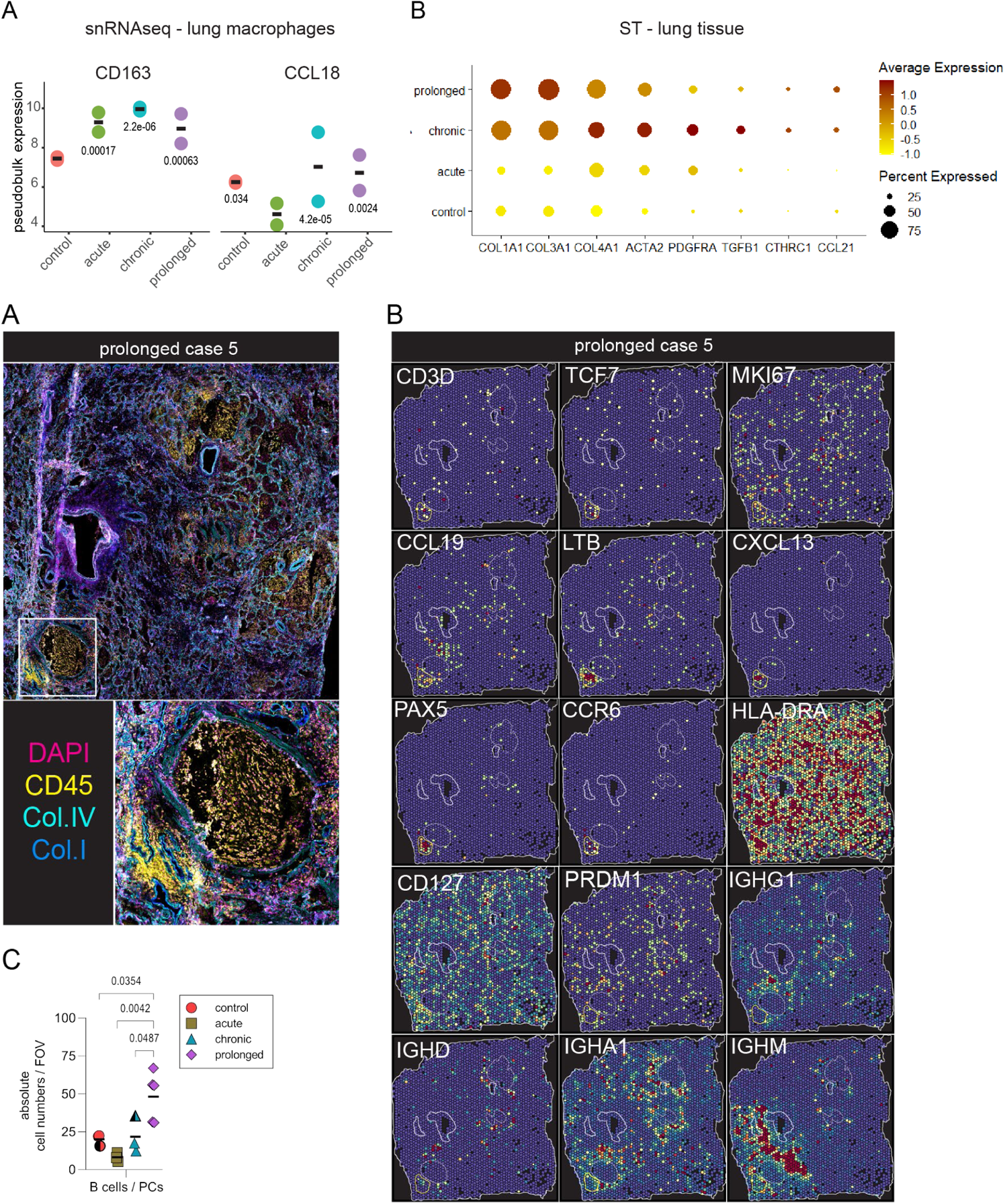
CCL21^+^ fibrovascular niches aim at the formation of tertiary lymphoid structures in prolonged COVID-19 lungs. (A) Immunofluorescence (IF) image depicting CD45 in yellow, DAPI in magenta, Collagen IV in cyan and Collagen I in blue. White square represents a region of interest (ROI), shown as an enlargement and depicting a highly infiltrated bronchus with a dense immune cell aggregate adjacent to it in the lower left corner. This tissue section of prolonged case 5 is adjacent to the one analyzed by ST and shown in (A) and in Fig. 4 A, B, right columns. (B) The spatial distribution of tertiary lymphoid structure-related transcripts is shown in a color-coded fashion as normalized enrichment scores (NES) in overlay with relevant tissue landmarks: intermediate-to-large vessels (thick white line), airway structures (dotted white line) and lymphoid structure (yellow line) (n = 2 lung samples). (C) Dot plot representation of the absolute counts of B cells / plasma cells per field of view analyzed by multiplex microscopy. Data (mean ± SD) are analyzed by ordinary one-way ANOVA with Tukey multiple comparisons test, where F = 8.469. (n = 32 FOVs)

Altogether, our data demonstrate that CCL21 production in fibrovascular niches eventually contributes to the formation of tertiary lymphoid structures in prolonged COVID-19 lungs.

## Discussion

Post-acute COVID-19 sequelae include various pulmonary and extra-pulmonary symptoms ^32^.They are not restricted to severe cases and represent a considerable burden on individuals post-infection, as well as on health care systems. Long-COVID syndrome is usually diagnosed if symptoms persist >2 months ^33^, but the mechanisms promoting and sustaining lung damage remain elusive. Recent reports highlighted the existing correlation between enriched cytotoxic lymphocytes and B cells within the airways of post-COVID-19 patients and impaired lung functionality ^11,12^. In this study, we stratify the samples based on disease duration and include a prolonged group (7 – 15 weeks of disease duration post-infection), in which active viral infection could neither be reported in the lungs nor in the LNs. Yet, those individuals showed persistent lung damage along with the increase in lymphocyte infiltration, as reported for Long-COVID previously ^11,12^. Thus, the prolonged group in this study temporally and symptomatically meets the Long-COVID criteria currently in use. Despite the non-negligible limitations of post-mortem studies, we think the mechanisms described here might also help to shed light into the pathophysiology of the Long-COVID syndrome.

By combining advanced microscopy approaches coupled to single-cell computational analysis with ST techniques in autopsy tissues, we delineated the fibrovascular niche, as a site where a dysregulated immune response leads to tissue repurposing in prolonged COVID-19 disease. In line with the idea that endothelial dysfunction is a key pathogenic mechanism in COVID-19 ^13,14^ and that endothelial cells are major participants in and regulators of inflammatory reactions ^34^, we identify EndMT as a mechanism driving fibrosis. When activated endothelial cells undergo EndMT, they are transcriptionally reprogrammed, their tight cellular junctions are disrupted and they turn into fibroblast-like cells. Consequently, they lose the expression of cell adhesion proteins, such as CD31, while mesenchyme-specific factors, including α-SMA, are upregulated ^35^. All these features are evident in our samples (Fig. 2A-C, Fig. 3A-C, Fig. S4C). In addition, we find that fibroblasts in COVID-19 lung fibrovascular niches express the chemokine receptor CCR8 (Fig. 3C). Its ligand, CCL18, has previously been linked to pulmonary inflammation and fibrosis ^36^ and it is the biomarker most consistently associated with negative outcomes in IPF ^37^. It was previously reported to be enriched in SARS-CoV-2 RNA^+^ myeloid cells in the lungs of COVID-19-deceased individuals ^3^. We have recently shown that alternatively activated, pro-fibrotic macrophages accumulate in fibrotic COVID-19 lungs ^10^. Here, we demonstrate that CD163^+^ macrophages accumulating in COVID-19 lungs produce CCL18, and spatially allocate this chemokine to the fibrotic patches (Fig. S5A, Fig. 3A). It remains to be evaluated whether the reported systemic increase in CCL18 during COVID-19 ^38^ is associated with fibrosis in other tissues beyond the lung, and whether increased CCL18 serum levels are associated to Long-COVID.

Our results also demonstrate an enrichment in *CCL21* in prolonged COVID-19 (Fig. S5B), being spatially restricted to the activated perivascular areas surrounding α-SMA-imprinted, intermediate-to-large vessels (Fig. 3A) and overlapping with EndMT-enriched areas (Fig. 3B). CCR7, the receptor for CCL21, is expressed by IPF fibroblasts in contrast to normal lung fibroblasts, and CCR7^+^ IPF-derived fibroblasts respond to CCL21 with activation, migration, survival, and proliferation ^39,40^. We show here that myofibroblasts in COVID-19 lungs also express CCR7 and, thus, can respond to the elevated CCL21 signals within the fibrovascular niches, particularly in the prolonged phases. On the other hand, although CCL21 is well known for its role recruiting CCR7^+^ T cells to secondary lymphoid organs to organize the adaptive immune response ^41,42^, CCL21 expression is also essential for the recruitment of effector T cells into the lungs ^26^. In line with that, our results show broad expression of CCR7 within the T cell population in the lungs of prolonged COVID-19, time-point in which absolute numbers of both CD4^+^ and CD8^+^ T cells are found significantly increased compared to all other disease groups analyzed (Fig. 4A, B). In addition, the marked upregulation of CD16 in both lung CD4^+^ and CD8^+^ T cell populations, together with Eomes and Granzyme B in CD4^+^ T cells (Fig. 4C and Fig. S6), suggests that T cells acquire NK cell-like functional properties and antibody-dependent cytotoxicity within the lung tissue, in line with published reports ^43^.

When comparing the T cell phenotype of the lung to that of the paired dLNs, our data point to a shift towards exhaustion within the inflammatory site, as opposed to a more conventional activated status in the secondary lymphoid organs (Fig. 4C, D and Fig. S6A-C). In line with the persistent T cell activation profile, we also report an increase in CD141^+^HLA-DR-DQ^+/−^ DCs in the LNs with disease progression, whereas these cells are undetectable in the lung (Fig. 4E, F and Fig. S3C). CD141^+^ DCs represent a unique myeloid DC subset that cross-presents viral Ag after their uptake of necrotic virus-infected cells ^44^. The lack of evidence for viral persistence in both lungs and dLNs from the prolonged group suggests that ongoing necrotic cell antigen cross-presentation in secondary lymphoid organs causes the persistent T cell activation. On the other hand, the lung T cell pool exhibits signs of exhaustion with broad and robust upregulation of PD1 in both, CD4^+^ and CD8^+^ T cell compartments, together with an increase in CD57 (Fig. 4C and Fig. S6A). These immunomodulatory molecules are co-expressed with the proliferation marker Ki67 (Fig. S6A). Exhausted T cells are emerging as a rather heterogeneous population with different functional properties and developmental potential, with TGF-β preserving an exhausted precursor population, which both limits and sustains long-term T cell responses in chronic inflammatory settings ^45^. We indeed report an increase of TGF-β in the lungs of COVID-19 chronic and prolonged groups at the tissue level (Fig. S5B), and previous studies have highlighted the essential role of TGF-β impacting on immune functions in COVID-19 disease ^31,46^.

Although the function of the exhausted population in the fibrovascular niches is unclear, we noticed co-expression of PD1 and ICOS, a phenotype linked to T follicular helper cells. Furthermore, the lymphoid accumulations show a strong upregulation of the CCR7 ligands *CCL19* and *CCL21* and *LTB* (Fig. 5B). These chemokines are essential for the formation of inducible bronchus-associated lymphoid tissue (iBALT) upon lung viral infection and inflammation ^47–49^. Additionally, lymphotoxin beta (LT-β) has been shown to further enhance CCL21 expression upon airway challenge, functioning as a positive feedback loop to attract additional CCR7^+^ effector T cells ^26^. The expression of the B cell attracting chemokine CXCL13 ^50^, another important factor in iBALT organization ^51^, is restrictively enhanced in the same lung area (Fig. 5B) and co-localizes with *PAX5,* along with evidence for a mucosal B cell profile, local B cell activation and antigen presentation. Thus, the data presented here strongly suggest the formation of iBALT in prolonged COVID-19 lungs, which contributes to the expansion of adaptive immune responses at sites of inflammation, thereby linking these tertiary lymphoid structures to lung pathology and fibrosis. Our data bring the activation of fibrovascular niches together with the generation of ectopic lymphoid follicles, consistent with their repurposing intoan inflammatory neo-tissue ^34^.

Our study proposes an inflammatory cascade of events causing an expansion of lung fibrovascular niches. Both CCL21/CCR7 and CCL18/CCR8 axis act synergistically upon the structural components within this niche to promote tissue remodeling. In addition, CCL21 attracts peripheral T cells into the inflammatory sites. Eventually, this culminates in the formation of tertiary lymphoid structures, further perpetuating chronic inflammation. This process, once triggered, occurs independently of active viral infection or latent persistence in lungs and dLNs, thereby presenting features of autoimmunity. Consequently, therapeutic strategies aiming at limiting viral replication in acute phases should be complemented with interventions interfering with this cascade, in order to avoid long-term tissue damage. This may be relevant not only for the treatment of severe COVID-19, but also have implications for the Long-COVID syndrome.

## Author contributions

A.E.H., H.R., A.P.R. and R.M. conceptualized the study. J.M, J.R, H.R. and F.H. organized the Biobank. M.L., C.D., S.E., G.A.H., B.O., J.I., F.H., P.E., H.S., T.A., J.S., V.C., L.E.S., M-F.M., T.C., A.H., H.R, R.N. and A.E.H. provided resources and expertise. G.M.G, J.L., A.L., R.M. and A.P.R. performed experiments. R.v.M, F.H, P.D., R.K., J.L., A.L., R.M. and A.P.R. analyzed the data. R.M., A.P.R., H.R. and A.E.H. interpreted the results and wrote the manuscript. H.S., T.A., B.O., S.B., R.K., R.M., A.P.R., M-F.M, R.N., H.R. and A.E.H. reviewed the manuscript.

## Methods

### Subject details

The lung and lung draining lymph node (dLN) samples included in this study have been collected in the Department of Pathology of Charité - Universitätsmedizin Berlin as part of the COVID-19 autopsy Biobank. This study was conducted in accordance with the declaration of Helsinki and with the approval of the Ethics Committee of the Charité (EA 1/144/927 13, EA2/066/20 and EA1/317/20) and the Charité - BIH COVID-19 research board. Autopsies were performed on the legal basis of §1 926 SRegG BE of the autopsy act of Berlin and §25(4) of the German Infection Protection Act. Autopsy consent was obtained from the families of the patients. Donor identities were encoded at the hospital before sharing for sample processing or data analysis. Some of the donors are part of the previously published cohorts in ^2,10,52^ and ^53^. All COVID-19 donors showed a positive result in real-time reverse transcription polymerase chain reaction (RT-PCR) in oropharyngeal swabs at the time of hospital admission. All relevant characteristics and clinical information of the donors is presented in Table 1.

### SARS-CoV-2-specific PCR

For assessment of SARS-CoV-2 RNA of the lung and dLN samples, unfixed and, where possible, non-cryopreserved (i.e., native) tissue samples were used. RNA was purified from ∼50 mg of homogenized tissue obtained from all samples by using the MagNAPure 96 system and the MagNAPure 96 DNA and Viral NA Large Volume kit (Roche) according to the manufacturer’s instructions. The controls without COVID-19 were validated by Rhonda PCR rapid COVID-19 test (Spindiag) according to the manufactures protocol on ∼50 mg of homogenized tissue together with a positive control.

Quantitative real-time PCR for SARS-CoV-2 was performed on RNA extracts with RT–qPCR targeting the SARS-CoV-2 E gene. Quantification of viral RNA was performed using photometrically quantified in vitro RNA transcripts as described previously ^54^. Total DNA was measured in all extracts by using the Qubit dsDNA HS Assay kit (Thermo Fisher Scientific). The RT–qPCR analysis was replicated at least once for each sample.

As a correlate of active virus replication in the tested tissues, subgenomic RNA (sgRNA) was assessed by using oligonucleotides targeting the leader transcriptional regulatory sequence and a region within the sgRNA encoding the SARS-CoV-2 E gene, as described previously ^55^. All information is presented in Table 1.

### Standard histological and immunohistochemical techniques

Lung FFPE tissue blocks were taken at the day of autopsy and fixed for 24 h in 4% paraformaldehyde at room temperature. Routine histological staining (HE, EvG and Prussian blue) was performed according to standard procedures.

Immunohistochemical for nucleocapsid staining was performed 1:4000 with #HS-452 011 (Mouse; Synaptic systems) on a Benchmark XT autostainer (Ventana Medical Systems) with standard antigen retrieval methods (CC1 buffer, pH 8.0, Ventana Medical Systems) using 4-μm-thick FFPE tissue sections. Immunohistochemistry sections were evaluated by at least two board-certified (neuro-)pathologists with concurrence. To biologically validate the immunohistological stainings, control tissues harboring or lacking the expected antigen were used. Staining patterns were compared to expected results as specified in the Fig. S2B, a semi-quantitative scoring was applied (0=no positive cell, +=single positive cells, ++=some positive cells, +++abundant positive cells and debris) (see Table 1 and Fig. S2B).

Fe3^+^ positive cells were scored using Prussian blue staining by two independent pathologists semi-quantitatively (0=no positive cell; +=single cells; ++=some cells and iron depositions; +++=abundant positive cells and iron depositions).

Images were analyzed using a Zeiss Axiolab 5 microscope.

### Electron microscopy

Frozen autopsy lung samples were thawn, fixed with 2.5% glutaraldehyde and 2% formaldehyde in 0.1 M sodium cacodylate buffer and embedded in Epon according to a routine protocol including en bloc staining with uranyl acetate and tannic acid ^52^. Large-scale digitization of ultrathin (70 nm) sections was performed using a Zeiss Gemini 300 field-emission scanning electron microscope in conjunction with a STEM detector via Atlas 5 software at a pixel size of 3–4 nm as previously described ^56^. Regions of interest from the large-scale datasets were saved by annotation (‘mapped’) and then recorded at very high resolution using a pixel size of 0.5–1 nm. Other images of virus particles in autopsy lung tissue of the same patient were previously published ^57–59^. We tried to optimize the sample handling for best preservation of the tissue, however effects mediated by autolysis and suboptimal fixation conditions cannot be excluded. We also analyzed two large-scale datasets of autopsy lung that we published previously ^52^ (see also www.nanotomy.org for open access of these two datasets).

### Confocal microscopy

Cryosections of lung tissue were cut at 7 mm on a MH560 microtome, transferred onto Superfrost Plus Gold slides (Fisher Scientific, Jena, Germany) and air dried. Sections were surrounded with a PAP pen (Zymed Laboratories, Jena, Germany) and rehydrated with PBS for 10 min. After that, sections were blocked for 20 min at room temperature with 10% goat serum in PBS. For immunofluorescence staining, antibodies were diluted in staining buffer (PBS, 5% BSA and 0.01%Triton) and incubated at room temperature for 60 min. The following antibodies were used for staining: anti-Collagen I-PE, anti-Collagen IV-FITC, anti-CD45-Alexa Fluor 647 and DAPI. After staining, samples were washed three times with PBS for 5 min and mounted with ProLong gold mounting medium (Life Technologies, Waltham, MA). Images were acquired by confocal microscopy using a Zeiss LSM 880 microscope with × 20/0.5 NA (air) objective at room temperature.

### MELC

#### Tissue preparation for MELC

Fresh frozen lung and dLN tissue was cut in 5 µm sections with a MH560 cryotome (ThermoFisher, Waltham, Massachusetts, USA) on cover slides (24 x 60 mm; Menzel-Gläser, Braunschweig, Germany) that had been coated with 3-aminopropyltriethoxysilane (APES). Samples were fixed for 10 minutes at room temperature (RT) with freshly opened, electron microscopy grade 2% paraformaldehyde (methanol- and RNAse-free; Electron Microscopy Sciences, Hatfield, Philadelphia, USA). After washing, samples were permeabilized with 0.2% Triton X-100 in PBS for 10 min at room temperature and unspecific binding was blocked with 10% goat serum and 1% BSA in PBS for at least 20 minutes. Afterwards, a fluid chamber holding 100 μl of PBS was created using “press-to-seal” silicone sheets (Life technologies, Carlsbad, California, USA; 1.0 mm thickness) with a circular cut-out (10 mm diameter), which was attached to the cover slip surrounding the sample. Prior to each MELC experiment fresh washing solution consistent of PBS with 5% MACS BSA (Miltenyi Biotec) and 0.02% Triton X-100 was prepared. The sample was placed on the sample holder and fixed with adhesive tape followed by accurate positioning of the binning lens, the light path, as well as Köhler illumination of the microscope.

#### MELC image acquisition

MELC image acquisition was performed as previously shown ^16,17^. In short, we generated the multiplexed histology data on a modified Toponome Image Cycler® MM3 (TIC) originally produced by MelTec GmbH & Co.KG Magdeburg, Germany ^60^. The ImageCycler is a robotic microscopic system with 3 main components:(1) an inverted widefield (epi)fluorescence microscope Leica DM IRE2 equipped with a CMOS camera and a motor-controlled XY-stage, (2) CAVRO XL3000 Pipette/Diluter (Tecan GmbH, Crailsheim, Germany), and (3) a software MelTec TIC-Control for controlling microscope and pipetting system and for synchronized image acquisition.

Each MELC experiment is a sequence of iterative cycles, each consisting of four steps: (i) pipetting of the fluorescence-coupled antibody onto the sample, incubation and subsequent washing; (ii) cross-correlation auto-focusing based on phase contrast images, followed by acquisition of the fluorescence images 3-D stack (+/− 5 z-steps); (iii) photo-bleaching of the fluorophore; and (iv) a second auto-focusing step followed by acquisition of a post-bleaching fluorescence image 3D stack (+/− 5 z-steps). In each four-step cycle up to four fluorescence-labeled antibodies were used, combining PE, FITC, APC and DAPI and images were acquired for two fields of view (FOV) in order to maximize area and cell numbers analyzed for each experiment and sample.

#### Antibody panel for MELC

The antibodies used are listed in the supplementary information table. The antibodies were stained in the indicated order.

#### MELC image pre-processing

Image pre-processing was conducted as previously described ^17^. In short, the reference phase-contrast image taken at the beginning of the measurement was used to align all images by cross-correlation. Afterwards, the signal of the bleaching image in each cycle and focal plane was used to subtract the background and correct the illumination of the fluorescence image obtained in the same cycle and focal plane ^60^.Thereby, tissue auto-fluorescence and potential residual signal from the previous cycle were removed. In case of uncoupled antibodies and to account for unspecific signal of the secondary antibody used, we subtracted the fluorescence image of the secondary antibody stained and acquired before the corresponding primary antibody, instead of the bleaching image. Then, an “Extended Depth of Field” algorithm was applied on the 3D fluorescence stack in each cycle ^61^. Images were then normalized in Fiji, where a rolling ball algorithm was used for background estimation, edges were removed (accounting for the maximum allowed shift during the autofocus procedure) and fluorescence intensities were scaled to the full intensity range (16 bit => 2^16^). The 2-D fluorescence images generated were subsequently segmented and analyzed.

#### Cell segmentation and single-cell feature extraction

Segmentation was performed in a two-step process as previously described ^17^, a signal-classification step using Ilastik 1.3.2 ^62^ and an object-recognition step using CellProfiler 3.1.8 ^63^. Ilastik was used to classify pixels into three classes: nuclei, membrane, and extracellular matrix (ECM). A probability map for each class was generated. Classification of images regarding membranes and ECM was performed by summing up a combination of images, using markers expressed in the respective compartments, while only the DAPI signal was used to classify nuclei. The random forest algorithm (machine-learning, Ilastik) was trained by manual pixel-classification in a small region of one data-set (approx. 6% of the image) and applied to the rest without re-training. CellProfiler was subsequently used to segment the nuclei and membrane probability maps and to generate nuclei and cellular binary masks, respectively. These masks were superimposed on the individual fluorescence images acquired for each marker, in order to extract single-cell information (mean fluorescence intensity, MFI) of each marker per segmented cell and their spatial coordinates.

#### MELC single-cell data transformation

Data was transformed using the hyperbolic arcsine function with a scale argument of 0.2.

#### Dimensionality reduction and clustering analysis

After exclusion of cells expressing none of the markers included in the MELC panel by setting a cut-off at MFI = 0.15, protein expression of 39.074 single cells from 14 lung samples and 32.258 single cells from 7 paired lung dLN samples was analyzed. Seurat package 4.0.0 ^64^ was used in R (R Core Team (2021, 2021) (https://www.R-project.org/) to perform mean centering and scaling, followed by principal component analysis (PCA), and reduced the dimensions of the data to the top 11 principal components in the lung and 10 principal components in the lung dLNs. Uniform Manifold Approximation and Projection (UMAP) was initialized in this PCA space to visualize the data on reduced UMAP dimensions. The cells were clustered on PCA space using the Shared Nearest Neighbor (SNN) algorithm implemented as *FindNeighbors* and *FindClusters* with *n.epochs = 500* and default parameters (*res = 0.8*) for the lung data set. We obtained 26 clusters that we merged to get relevant populations for our analysis based on canonical lineage markers. We ended up with 8 cell clusters that were manually annotated based on cell-type-specific markers found to be differentially expressed. Protein expression of the main cell clusters was visualized using *UMMAPPlot, DotPlot* and *VlnPlot* functions from Seurat. 7 cell clusters in the lung dLN data set were obtained and visualized using the same algorithms and functions, but with *res = 0.2*.

#### Niche analysis

We compared the phenotype of the CD4^+^ and CD8^+^ T cell clusters located in the fibrovascular niche to the ones located in the epithelial niche. We defined the niche as the area of a 32 pixel-radius (equivalent to 10 µm; the average size of a hematopoietic cell) around each endothelial or epithelial cell, similar to previous analysis ^17^. We extracted the MFI of each T cell-related marker for the sum of all fibrovascular niches and epithelial niches and displayed the expression profile as heat map showing the z-score values.

### Spatial transcriptomics

Visualisation of gene expression in lung tissue was performed using 10x Visium spatial gene expression kit (10x Genomics) following manufacturers protocol. The four capture areas in a 10x Genomics Visium Gene Expression slide consist of 5000 spots with DNA oligos for mRNA capture that have a unique spatial barcode and a Unique Molecular Identifier (UMI). Each spot has 55 µm diameter and can therefore capture mRNA from 1 to 10 cells. Tissue sections from the same fresh frozen human lung samples used for MELC and snRNA-seq were analyzed by ST. Briefly, control (n=3) and COVID-19 lung samples (n=9) from donors categorized based on disease durations (acute n=3, chronic n=3, prolonged n=3) were cut into 10μm sections using a MH560 cryotome (ThermoFisher, Waltham, Massachusetts, USA), and mounted on 10X Visium slides, which were pre-cooled to −20°C. The sections were fixed in pre-chilled methanol for 30 min, stained with CD45-AF647, CD31-AF594 and DAPI for 30min and imaged using an LSM 880 confocal microscope (Zeiss). The sections were then permeabilised for 10 minutes and spatially tagged cDNA libraries were constructed using the 10x Genomics Visium Spatial Gene Expression 3’ Library Construction V1 Kit. cDNA libraries were sequenced on an Illumina NextSeq 500/550 using 150 cycle high output kits with sequencing depth of ∼5000 reads per spot. Frames around the capture area on the Visium slide were aligned manually and spots covering the tissue were manually selected based on the immunofluorescence staining, using Loupe Browser 5.1.0 software (10x Genomics). Sequencing data was mapped to GRCH38-2020-A reference transcriptome using the Space Ranger software (version 1.3.0, 10x genomics) to derive a feature spot-barcode expression matrix.

Space Ranger outputs (feature barcode matrices and image data) from 31.801 barcoded spatial spots from twelve 10x Visium capture areas were filtered and integrated in R (version 4.1.0) with Seurat (version 4.0.4). Spots with at least 250 detected genes and less than 13% mitochondrial content were combined from each library. For that, SCTransform normalization was performed ^66^, in order to harmonize the Pearson residuals from each RNAseq data set. We selected *nfeatures = 3000* for downstream integration. Then, principal component analysis (PCA) was performed on the matrix composed of spots and gene expression (UMI) counts. The dimensions of the data were thereby reduced to the top 50 principal components. Uniform Manifold Approximation and Projection (UMAP) was initialized in this PCA space to visualize the data on reduced UMAP dimensions. The spots were clustered on PCA space using the Shared Nearest Neighbor (SNN) algorithm implemented as *FindNeighbors* and *FindClusters* with parameters *k =30* and *res = 0.3*. The method returned 12 spot clusters that represented lung microanatomical areas with distinct transcriptional signatures and that were then visualized on the UMAP space.

The integrated data was also used to perform Gene Set Enrichment Analysis (GSEA) with the *fgsea* package (version 1.18.0) ^67^, which compared the Wilcoxon-ranked genes from each data set to the Hallmark, Reactome and Gene Ontology gene sets from MSigDB (v7.4). Only significantly enriched genes (adjusted *p* < 0.05) were taken into account to measure the Normalized Enrichment Score (NES). We then performed single sample gene set enrichment analysis (ssGSEA) with the *escape* package (version 1.2.0) ^68^, where the normalized enrichment scores can be visualized on the spatial feature plot in Seurat. NES range was clipped to the first and last five percentile to enhance visibility.

### Single-nucleus RNA-sequencing

#### Single-cell isolation and library preparation

Each lung tissue sample was minced and placed in digestion medium (500 U/ml Collagenase, I.5 U/ml Dispase and 1 U/ml DNAse) for 1 h at 37°C. Cells were filtered through a 70 μm strainer and enzymatic reaction was stopped by cold RPMI with 10 % fetal bovine serum and 1 % L-Glutamine. Cells were washed with 50 ml cold RPMI with 10 % fetal bovine serum and 1 % L-Glutamine and red blood cells were lysed using Red blood cell lysis solution (MiltenyiBiotec). Finally, cells were filtered using a 40 μm FlowmiR Cell Strainer (Millipore) and re-suspended in PBS supplemented with 2 % fetal bovine serum at the concentration of 10.000 cells/μl for scRNA-Seq. The single-cell capturing and downstream library constructions were performed using the Chromium Single Cell 3ʹ V3.1 library preparation kit according to the manufacturer’s protocol (10x Genomics). Full-length cDNA along with cell-barcode identifiers were PCR-amplified and sequencing libraries were prepared. The constructed libraries were either sequenced on the Nextseq 500 using 28 cycles for read 1.55 cycles for read 2, and 8 index cycles, or on the Novaseq 6000 S1 using 28 cycles for read 1, 64 cycles for read 2, and 8 index cycles, to a median depth of 36000 reads per cell.

#### Single-cell RNA sequencing and analysis

The Cell Ranger Software Suite (Version 3.1.0) was used to process raw sequencing data with the GRCh38 reference. Single-cell RNA sequencing data analysis was performed in R (version 3.6.1) with Seurat (version 3.1.1). Cells with at least 500 and less than 5000 detected genes and less than 10% mitochondrial content were combined from each library. Library depth (total number of UMIs) was regressed out when scaling data and libraries from different donors were integrated using *IntegrateData*. Cluster annotation was performed using the Human Lung Cell Atlas reference dataset ^69^ and Seurat’s *TransferData* workflow as published elsewhere ^19^.

### Light sheet microscopy

#### Whole-mount staining and optical clearing of human lung tissue

About 3 mm PFA-fixed human lung cubic samples were processed for tissue permeabilization and decolorization based on a recently published approach ^70^. All steps were performed permanently shaken on a tube rotator (MACSmix^TM^, Miltenyi Biotec). We incubated the samples first in 5mL 25% w/v Quadrol solution (N,N,N′,N′-Tetrakis-(2-hydroxypropyl)-ethylendiamin, Sigma-Aldrich) for 2 days at 37°C, followed by 5mL 25% w/v Quadrol and 10% w/v CHAPS solution ((3-[3-cholamidopropyl)dimethylammonio]-1-1propanesulfonate, Sigma-Aldrich) for 5 days at 37°C ^70^. After initial decolorization and permeabilization, we followed the SHIELD protocol for PFA-fixed human samples ^71^. Therefore, samples were incubated in SHIELD-OFF solution at 4 °C for 3 days and then placed in SHIELD-ON Buffer and SHIELD-Epoxy Solution in a ratio of 1:1 at RT for 24h. SHIELD preservation is then completed and we washed the samples for 1 day in PBS with three times refresh at 4°C. Next, we actively cleared the samples for 3 days using stochastic electrotransport (SmartClear Pro, LifeCanvas Technologies) and performed the same washing step as before.

For immunostaining, lung samples were incubated in blocking buffer (0.2% Triton X-100/10% DMSO/5% FCS/PBS) at 37 °C for 1 day and then in antibody incubation buffer (0,5% FCS/3% DMSO/0.2%Tween-20/PBS) at 37°C for 7 days containing the respective antibodies (as indicated for the respective samples). We used following antibodies and dilutions for staining: Sytox-AF488 (1:40.000, Thermo Fisher), ER-TR7-AF546 (1^st^ 1:200/2^nd^ 1:100, Santa Cruz Biotechnology) or near-infrared (NIR) CD3-iFluor790 (1:50, AAT Bioquest, Inc., Sunnyvale, CA, USA). After staining samples were washed first with washing buffer (0.2%Tween-20/PBS) for 1 day at 37°C and with PBS at 4°C with three times refresh each. Then, immunolabeled samples were incubated for index matching first in 50% EasyIndex (LifeCanvas Technologies, refractive index –RI-1,456) in ddH_2_O for 1 day and second in 100% Easy Index until transparency was reached within 1-2 days.

#### Sample mounting and light-sheet microscopy imaging

We slightly glued the human lung samples on a plastic plate and immersed the sample in 100% Easy Index solution in a 5×5×45mm quartz cuvette (msscientific Chromatographie-Handel GmbH). For image acquisition, we immersed the smaller cuvette containing the lung sample in the bigger quartz cuvette (LaVision Biotec), filled with fused silica matching liquid with a RI of 1,459 (Cargille Laboratories). To acquire light-sheet images we used an Ultramicroscope II (LaVision BioTec) coupled to an Olympus MVX10 zoom body providing a zoom ratio from 0.63x - 6.3x and a 2x dipping objective (Olympus MVPLAPO2XC/0.5 NA [WD= 5.6mm, corrected]) resulting in a total magnification ranging from 1.36x - 13.56x. The Ultramicroscope II featuring a lateral resolution of ∼4 µm was equipped with an Andor Neo sCMOS Camera with a pixel size of 6.5x 6.5 µm² and a LaVision BioTec Laser Module featuring the following filter sets: ex 488 nm, em 520/50 nm; ex 561 nm, em 620/60 nm; ex 785 nm, em 835/70 nm. We used the 488 nm ex for generation of autofluorescent signals and detection of nuclear Sytox-AF488 signal, the 561 nm ex for detection of ER-TR7-AF546 signal and the NIR 785 nm ex for detection of CD3-iF790. We imaged the longest wavelength first to avoid photobleaching during imaging and performed a linear z-adaption of the ex. 785 nm laser. We used a total optical zoom of 8,61x and a z-step of 5 µm for all acquisitions. Bigger tile scans were acquired using 10% overlap along longitudinal x-axis and y-axis of the human lung sample and the laser power was adjusted depending on the intensity of the signal (in order to not reach the saturation.

#### Image processing

We separately acquired 16-bit grayscale TIFF images for each channel by light sheet microscopy with the ImSpector software (LaVision Biotec). Tiff stacks were converted (ImarisConverterx64) into Imaris files (.ims) and stitched by Imaris Stitcher. We used Imaris (Bitplane) for 3-D and 2-D image visualization, snapshot creation and video generation. We performed background subtraction in accordance with the diameter of the respective structures to eliminate unspecific background signals.

### Quantification and statistical analysis

Statistical analysis was performed with GraphPad Prism® 9.2.0 (Graph Pad Software, LA Jolla, CA, USA). The tests used and the exact *p* values are described within each figure and figure legend. Significance was defined by *p* < 0.05.

## Data and code availability

De-identified human/patient spatial transcriptomics data have been deposited at Gene Expression Omnibus (https://www.ncbi.nlm.nih.gov/geo/), under record GSE190732, and are available for reviewers. Microscopy data, as well as any additional information required to re-analyze the data reported in this paper will be shared by the lead contact upon request. This study did not use any original code.

## Data Availability

De-identified human/patient spatial transcriptomics data have been deposited at Gene Expression Omnibus (https://www.ncbi.nlm.nih.gov/geo/), under record GSE190732, and are available for reviewers. Microscopy data, as well as any additional information required to re-analyze the data reported in this paper will be shared by the lead contact upon reasonable request.

## Acknowledgements

The authors are most grateful to the patients and their relatives for consenting to autopsy and subsequent research. We thank Lars Philipsen for his assistance with multiplex microscopy and Gudrun Holland for her support in acquisition of the electron microscopy data. The authors are indebted to Ralf Uecker for his valuable technical support. We thank Daniel Wendisch for fruitful discussions.

This study was supported by funding from the Deutsche Forschungsgemeinschaft, HA5354/10-1, SPP1937 (HA5354/8-2) and TRR130 P17 to A.E.H., TRR 130 C01 and SFB 1444 P14 (to A.E.H. and R.N.). R.A.N. was supported by DFG 1167/5-1. H.R. was supported by DFG (RA 2491/1-1 and CRC 130 TP17). A.H. was supported by Berlin University Alliance GC2 Global Health (Corona Virus Pre-Exploration Project), BMBF (RAPID and Organo-Strat, alvBarriere-COVID-19) as well as DFG (SFB-TR 84, B6 / Z1a), by the Berlin Institute of Health (BIH), Charité 3 R, and Charité-Zeiss MultiDim. S.E., F.H., H.R. and R.M. were supported by BMBF (Defeat Pandemics, Organo-Strat), F.H. was supported by DFG under Germany’s Excellence Strategy EXC-2049-390688087, as well as SFB TRR 167 and HE 3130/6-1. B.O. was funded through the BIH Clinical Single Cell Bioinformatics Pipeline. Furthermore, we thank the Charité Foundation (Max Rubner Preis 2016) for financial support to C.D.

## Extended data

**Figure S1.**
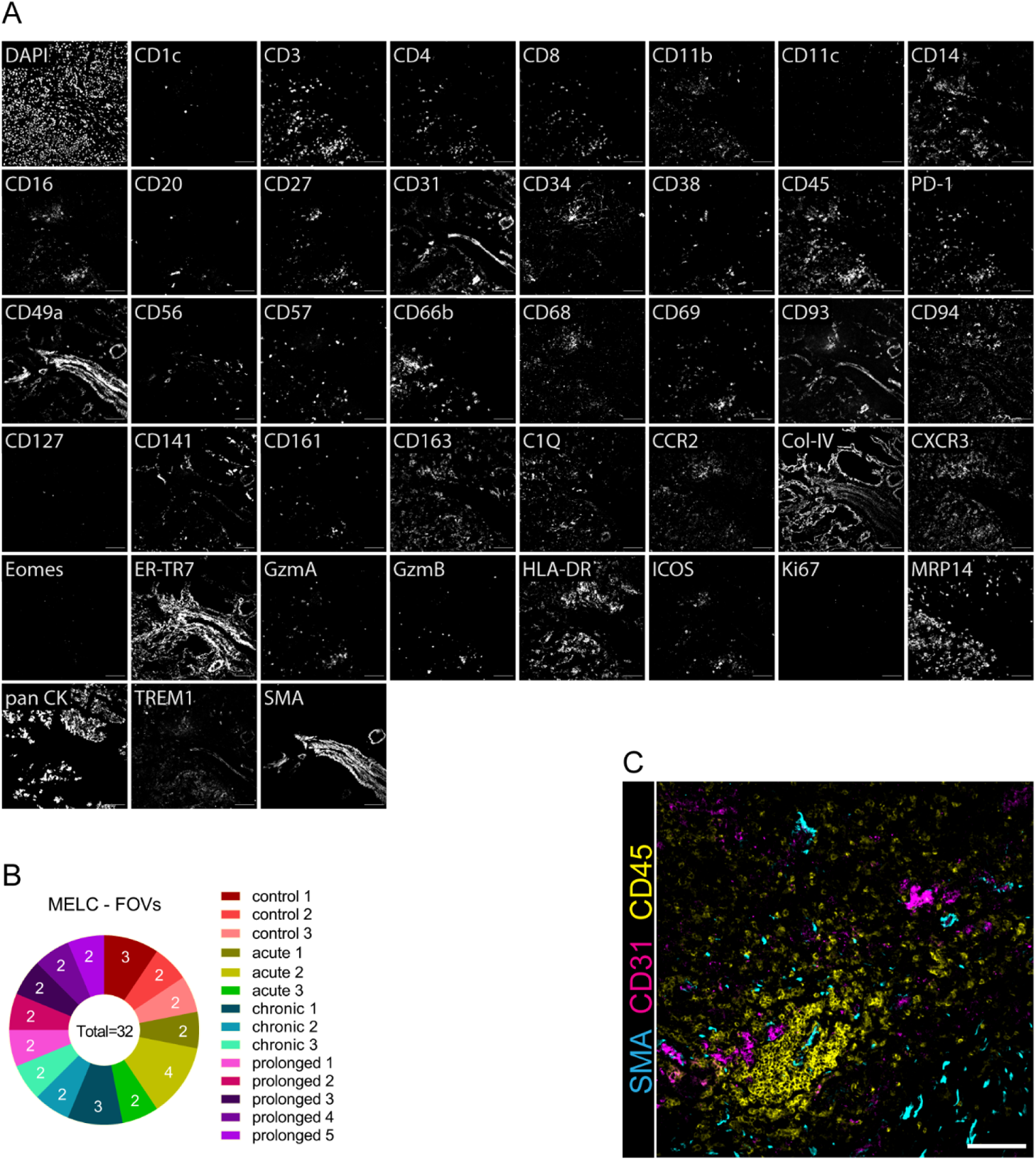
Overview of markers and samples analyzed by multiplex microscopy. (A) Overview of 43-marker MELC panel in human lung. Each image depicts the same field of view (FOV), sequentially stained with the depicted fluorescence-labelled antibodies. Images contain 2048 x 2048 pixels and are generated using an inverted wide-field fluorescence microscope with a 20x objective, a lateral resolution of 325 nm and an axial resolution above 5 µm. (B) Overview of the FOVs acquired for each donor by multiplex microscopy. (C) Immunofluorescence overlay of αSMA (cyan), CD45 (yellow) and CD31 (magenta) acquired by multiplex microscopy in the lungs of prolonged case 4. Scale bar: 100 µm. (n=32)

**Figure S2.**
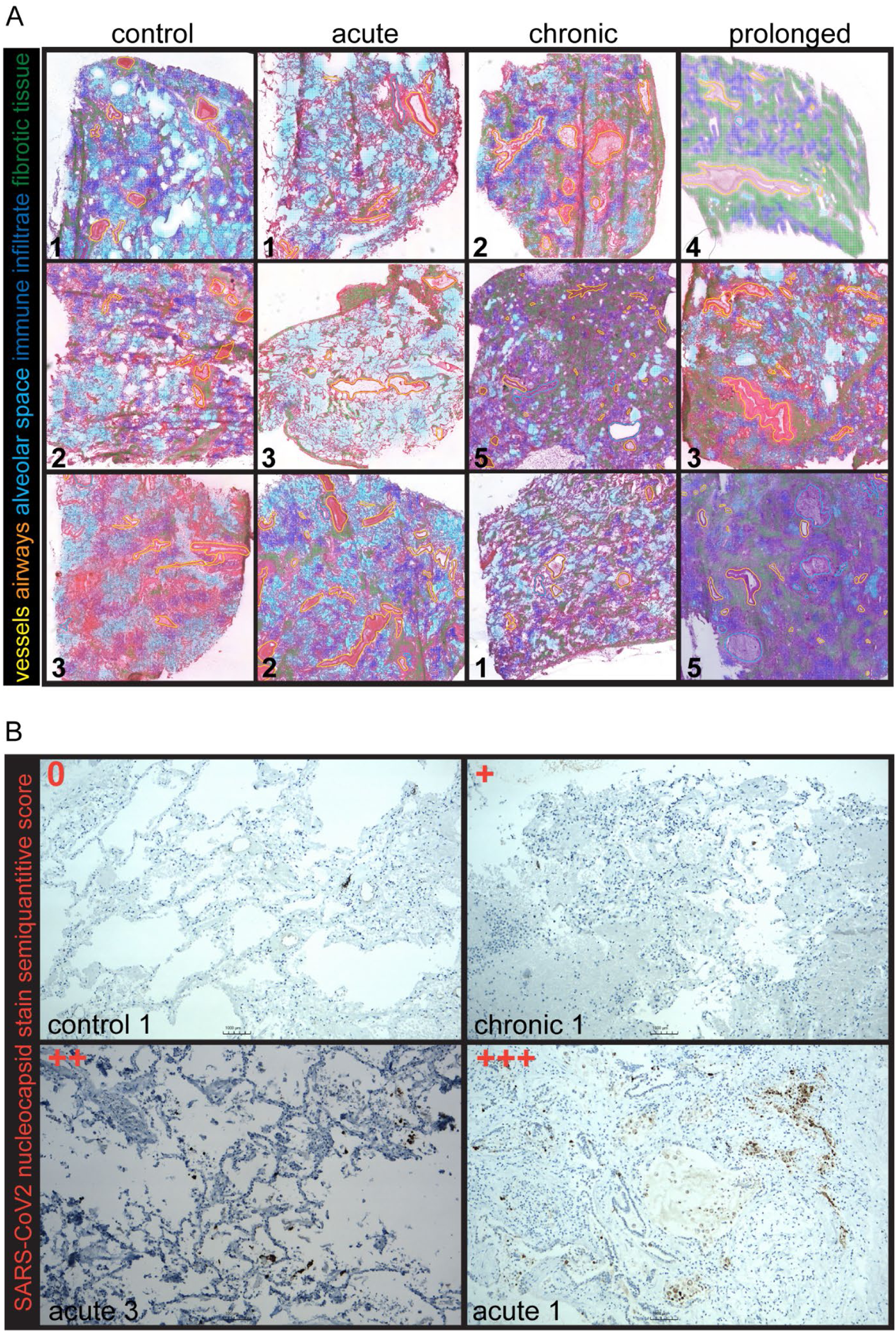
Histopathological evaluation of lung samples from COVID-19 donors. (A) HE staining of human lung tissue from several non-COVID-related pneumonia donors, as well as from COVID-19 cases was used by a blinded and trained pathologist to assess histopathological changes and to annotate recognizable lung structures to be used as landmarks for further analyses. Vessels are marked in yellow lines, airways in cyan lines, alveolar space in cyan areas, highly infiltrated areas in blue areas and fibrotic areas in green areas. Numbers represent donor IDs, as shown in Table 1. (n = 12 lung samples). See also Fig. 1, 2, 4 and 6. (B) 4 representative images illustrating semiquantitative scoring of SARS-CoV-2 nucleocapsid immunohistochemical staining.

**Figure S3.**
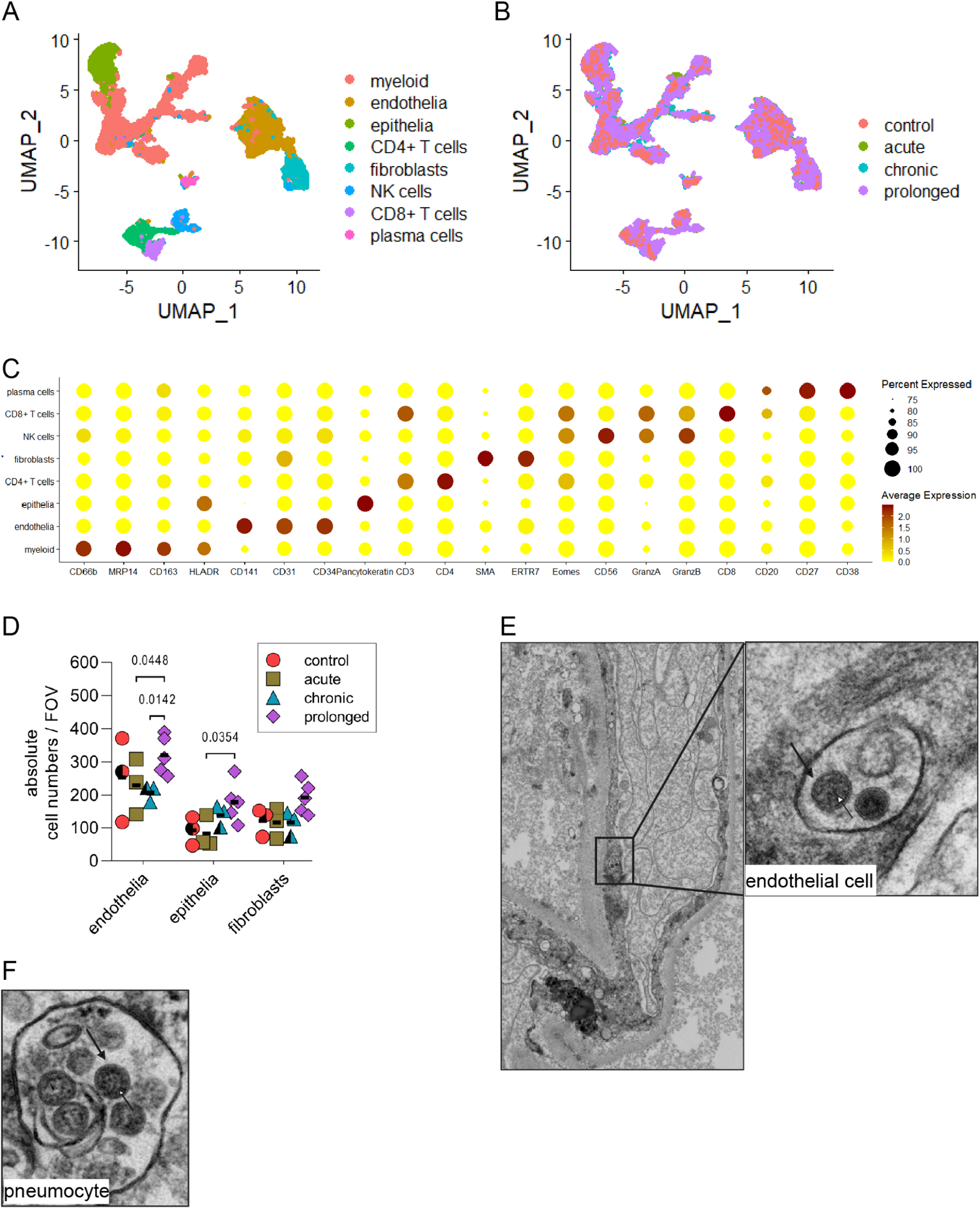
Dimensionality reduction and clustering analysis of multiplex microscopy single-cell data and ultrastructural findings in autopsy lung tissue with two putative coronavirus particles within an endothelial cell. (A, B) Uniform Manifold Approximation and Projection (UMAP) of segmented single-cell multiplex microscopy data from 14 lung samples showing 8 clusters. (A) Color code depicts the annotated cell clusters, based on lineage-defining markers as shown in (C). (B) Color code represents the disease group. (C) Dot plot showing the expression profile of the 8 cell clusters identified by multiplex microscopy and as defined in (A). (D) Dot plot of the absolute numbers of endothelial and epithelial cells as well as fibroblasts within the different cohort populations shows significance in altered cell numbers in the endothelial and epithelial compartment in later disease phase of COVID-19 infection. Data (mean ± SD) are analyzed by two-way ANOVA with Fisher’s LSD test, where DF = 2 and F = 19.05 (n = 32 FOVs). (E-F) Ultrastructural images of an acute lung tissue sample showing two putative coronavirus particles within a membrane compartment in an endothelial cell (E; magnified). The coronavirus particles within pneumocytes (F) can be unequivocally identified as coronavirus particles due to their larger number and, thus, as a collective clearly fulfil all morphologic criteria: faint but detectable surface projections (F; black arrow) and an electron dense, partly granular interior due to ribonucleoprotein (RNP, e.g. F; white arrow). In contrast, the two particles in (E) cannot unequivocally be identified as coronavirus particles, albeit they fulfill some morphologic criteria (surface projections; black arrow, possible RNP; white arrow), as we only detected these two examples in endothelial cells of six entirely digitized and analyzed ultrathin sections of autopsy lung of three different patients.

**Figure S4.**
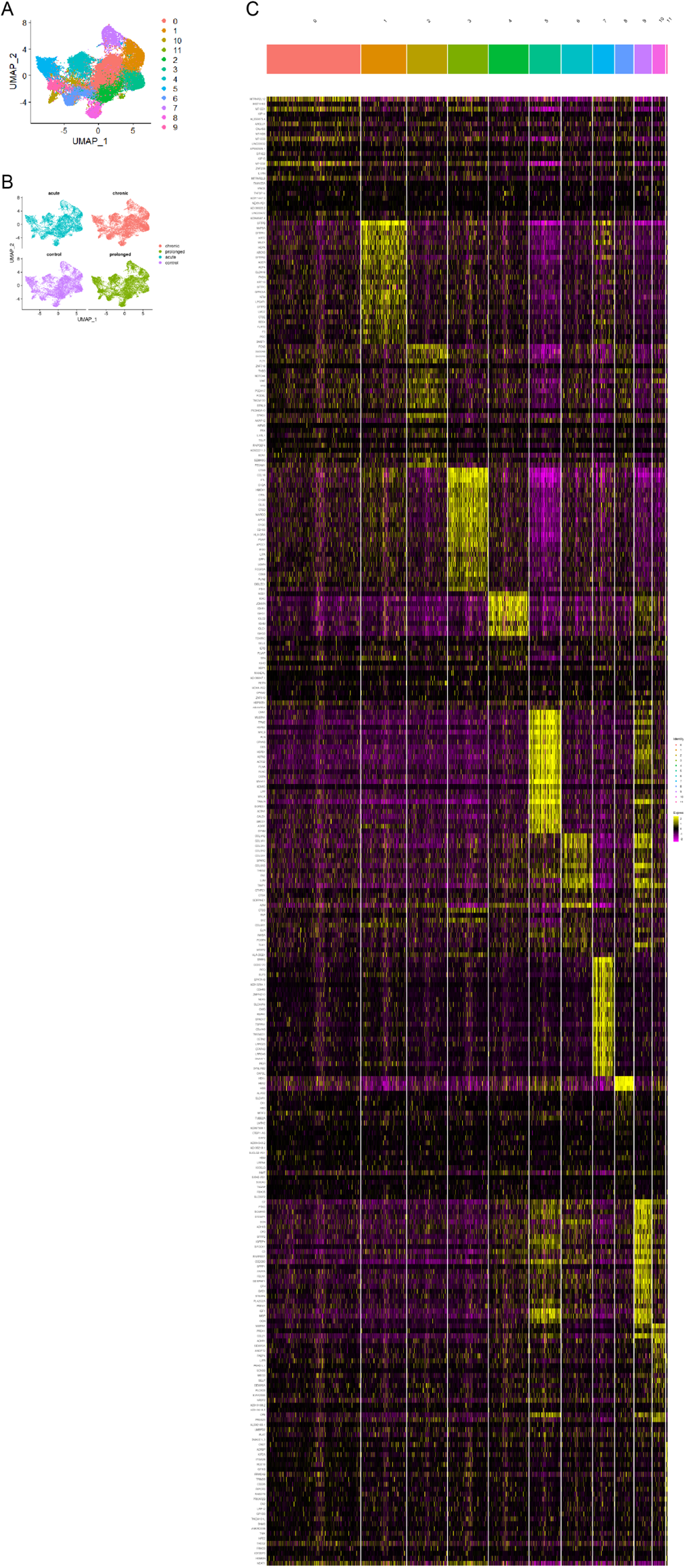
Dimensionality reduction and cluster analysis of ST data. ST feature spot-barcode expression matrix was filtered and integrated in R with Seurat (see methods). (A) Uniform Manifold Approximation and Projection (UMAP) of spatial transcriptomics data from 12 lung samples showing 12 clusters. (A) Color code depicts 12 unsupervised spot clusters. (B) Color code represents the disease group. (C) Heatmap showing the top 25 differentially expressed genes for each cluster depicted in (A). (n = 12 lung tissue sections).

**Figure S5.**
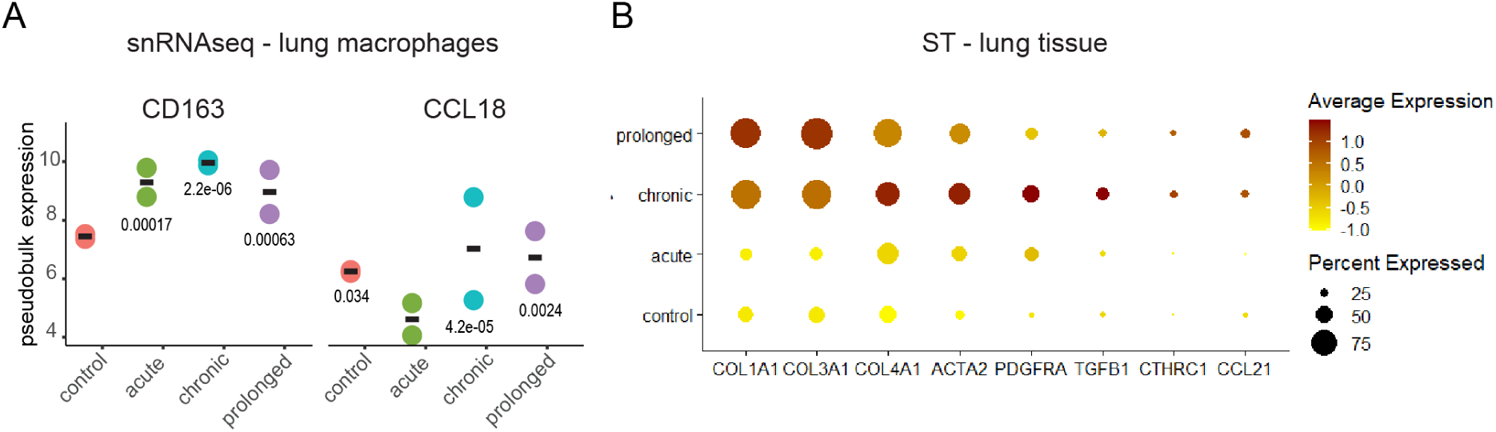
ST analysis reveals an increasing fibrotic signature in COVID-19 lungs along with disease progression. (A) Dot plot of *CD163* and *CCL18* pseudobulk expression levels within the macrophage population of each disease group as analyzed by single nuclei RNA sequencing, as analyzed in ^19^. Data (mean ± SD) are analyzed by Wald test using DESeq2 on aggregated counts, where DF=4 (n = 8 tissue samples). (B) Dot plot depicting the normalized enrichment scores (NES) and leading edge counts (LEC) of fibrosis-related genes in each disease group analyzed by spatial transcriptomics (ST). (n = 12 lung tissue sections).

**Figure S6.**
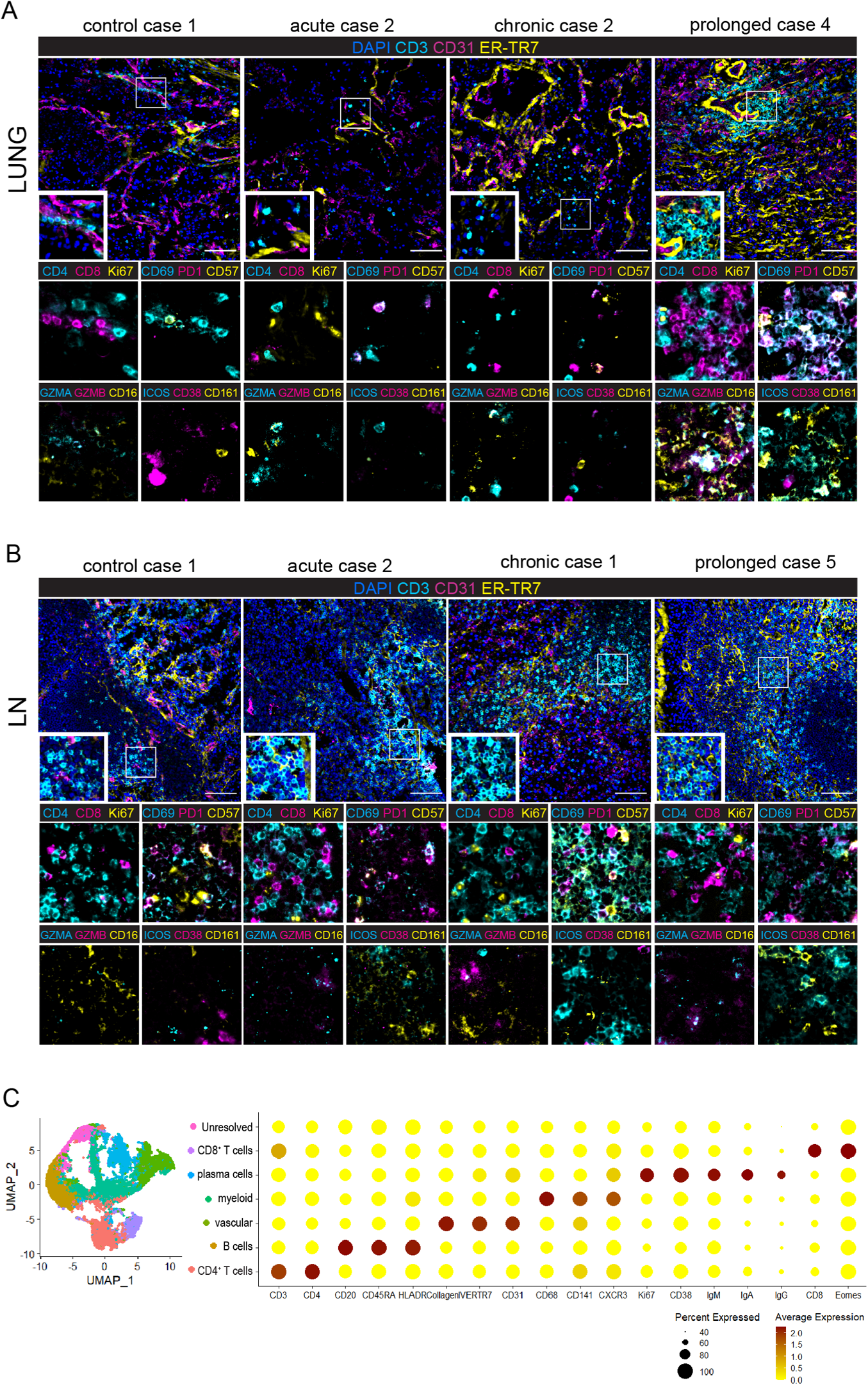
Non-conventional T cells accumulate in fibrovascular niches of prolonged COVID-19 lungs, while T cells in the draining lymph nodes show a conventional activated profile. First row: representative multiplex microscopy images from each disease group showing nuclear staining DAPI (blue), the fibroblast marker ER-TR7 (yellow), the endothelial marker CD31 (magenta) and the T cell marker CD3 (cyan). White squares represent regions of interest (ROIs) shown as enlargements. For each lung area the identical ROI is depicted below in 4 overlays of different markers. Upper left corner of the enlargements: Ki67 (yellow), CD4 (cyan) and CD8 (magenta). Upper right corner of the enlargements: CD69 (cyan), PD1 (magenta) and CD57 (yellow). Lower left corner of the enlargements: Granzyme A (cyan) and B (magenta) and CD16 (yellow). Lower right corner of the enlargements: CD38 (magenta), CD161 (yellow) and ICOS (cyan). (n = 32). (B) The same arrays of overlays as in (A) are shown in the draining lymph nodes. (n = 9). (C) Left side: Uniform Manifold Approximation and Projection (UMAP) of segmented single-cell multiplex microscopy data from 7 lymph node samples showing 7 clusters. Color code depicts the annotated cell clusters, based on lineage-defining markers as shown in the dot plot (right side) showing the expression profile of each of the 7 cell clusters identified by multiplex microscopy.

**Figure S7.**
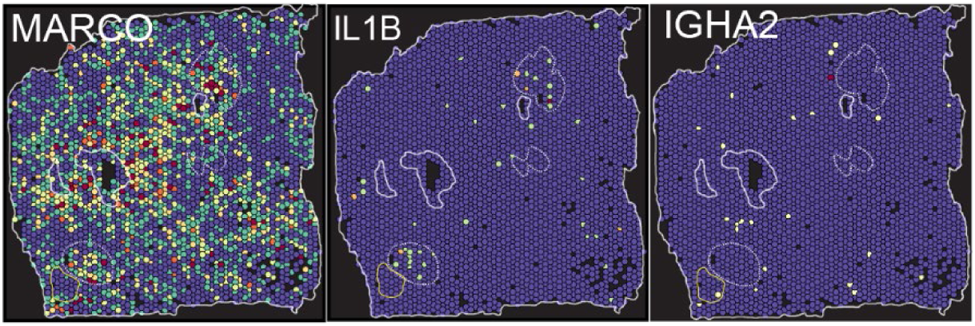
Spatial distribution of additional transcripts. Color-coded tissue spots analyzed by ST depict the tissue distribution of the normalized enrichment score (NES) of *MARCO*, *IL1B* and *IGHA2* within the same lung section from prolonged case 5 as shown in Fig. 6. (n = 12 lung tissue sections).

Video S1. LSFM of COVID-19 lung from acute case 3 showing a thrombus attached to the vessel wall. Yellow color represents ER-TR7 staining, signals shown in magenta result from autofluorescence.

Video S2. LSFM of COVID-19 lung from acute case 3 stained with antibodies against ER-TR7 for visualization of the fibroblast network.

Video S3. LSFM of COVID-19 lung from prolonged case 5 stained with anti-ER-TR7 for visualization of the fibroblast network.

Video S4. LSFM of COVID-19 lung from acute case 3 stained with sytox green (nuclei) in magenta and anti-CD3 in cyan.

Video S5. LSFM of COVID-19 lung from prolonged case 5 stained with sytox green (nuclei) in magenta and anti-CD3 in cyan.

