## Supplementary Table 1 for "Local CCL18 and CCL21 expand lung fibrovascular niches and recruit lymphocytes, leading to tertiary lymphoid structure formation in prolonged COVID-19"

### Supplementary information

Table S1: Reagents and Resources

| REAGENT or RESOURCE | SOURCE | IDENTIFIER |
| --- | --- | --- |
| **Antibodies** | | |
| DAPI | Roche | Cat# 10236276001, N/A |
| Fibronectin | Thermo Fisher | Cat# PA5-29578; RRID:AB_2547054 |
| Rabbit IgG-PE | Rockland | Cat# 711-708-127, RRID:AB_218957 |
| CCR2-PE | Miltenyi Biotec | Cat# 130-118-338, RRID:AB_2751486 |
| CD163-PE | Biolegend | Cat# 333605, RRID:AB_1134005 |
| CD56-PE | Miltenyi Biotec | Cat# 130-098-137, RRID:AB_2661200 |
| CD1c-PE | Miltenyi Biotec | Cat# 130-113-864, RRID:AB_2726358 |
| CD3-PE | Miltenyi Biotec | Cat# 130-113-139, RRID:AB_2725967 |
| CD14-PE | Miltenyi Biotec | Cat# 130-113-709, RRID:AB_2726250 |
| Eomes-PE | Thermo Fisher | Cat# 14-4877-80, RRID:AB_2572881 |
| CD45-PE | Miltenyi Biotec | Cat# 130-113-118, RRID:AB_2725946 |
| CXCR3-PE | Miltenyi Biotec | Cat# 130-101-379, RRID:AB_2655734 |
| PD1-PE | Miltenyi Biotec | Cat# 130-120-388, RRID:AB_2752074 |
| CD16-PE | Miltenyi Biotec | Cat# 130-113-955, RRID:AB_2726428 |
| CD93-PE | Miltenyi Biotec | Cat# 130-098-436, RRID:AB_2659615 |
| CD4-PE | Miltenyi Biotec | Cat# 130-113-214, RRID:AB_2726025 |
| Granzyme A-PE | Miltenyi Biotec | Cat# 130-123-973, RRID:AB_2889678 |
| CD31-PE | R and D Systems | Cat# FAB3567P, RRID:AB_2279388 |
| ICOS-PE | Miltenyi Biotec | Cat# 130-120-155, RRID:AB_2784102 |
| TREM1-PE | Miltenyi Biotec | Cat# 130-101-033, RRID:AB_2657706 |
| CD20-PE | Miltenyi Biotec | Cat# 130-113-374, RRID:AB_2726143 |
| CD11b-PE | Miltenyi Biotec | Cat# 130-110-553, RRID:AB_2654665 |
| CD8-PE | Miltenyi Biotec | Cat# 130-113-720, RRID:AB_2726261 |
| CD68-PE | Miltenyi Biotec | Cat# 130-118-486, RRID:AB_2784270 |
| CD127-PE | Miltenyi Biotec | Cat# 130-113-414, RRID:AB_2733759 |
| CD11c-PE | Miltenyi Biotec | Cat# 130-113-580, RRID:AB_2726180 |
| Granzyme B-PE | Miltenyi Biotec | Cat# 130-116-654, RRID:AB_2727639 |
| CD69-PE | Miltenyi Biotec | Cat# 130-112-613, RRID:AB_2659065 |
| CD94-PE | Miltenyi Biotec | Cat# 130-098-973, RRID:AB_2659624 |
| CD141-PE | Miltenyi Biotec | Cat# 130-114-188, RRID:AB_2751233 |
| CD27-PE | Miltenyi Biotec | Cat# 130-114-166, RRID:AB_2726471 |
| CD38-PE | Miltenyi Biotec | Cat# 130-113-427, RRID:AB_2733813 |
| CD57-PE | Miltenyi Biotec | Cat# 130-111-963, RRID:AB_2658747 |
| CD161-PE | Miltenyi Biotec | Cat# 130-114-119, RRID:AB_2733771 |
| HLA-DR,DP,DQ-PE | Miltenyi Biotec | Cat# 130-120-715, RRID:AB_2752176 |
| CD34-PE | Miltenyi Biotec | Cat# 130-113-741, RRID:AB_2726281 |
| CD66b-PE | Miltenyi Biotec | Cat# 130-122-966, RRID:AB_2811418 |
| Pancytokeratin-PE | Arigo Biolaboratories | Cat# ARG56130 |
| Ki67-FITC | Dako | F268 |
| CD49a-PE | Miltenyi Biotec | Cat# 328304, RRID:AB_1236407 |
| Collagen IV-FITC | Antibodies-Online | Cat# ABIN376119, RRID:AB_10763557 |
| ER-TR7-PE | Thermo Fisher | Cat# MA1-40076, RRID:AB_1074409 |
| SMA-FITC | Abcam | Cat# ab8211, RRID:AB_306359 |
| CD45RA-PE | Miltenyi Biotec | Cat# 130-113-366, RRID:AB_2726136 |
| C1q-FITC | DAKO | Cat# F0254, RRID:AB_2335713 |
| MRP14-PE | Miltenyi Biotec | Cat# 130-114-516, RRID:AB_2726684 |
| Collagen I-PE | Biolegend | Cat# 303126, RRID:AB_2563303 |
| IgA-PE | Miltenyi Biotec | Cat# 130-114-002  RRID:AB_2733860 |
| IgA2-PE | Miltenyi Biotec | Cat# 130-117-874  RRID:AB_2728061 |
| IgM-PE | Miltenyi Biotec | Cat# 130-122-930  RRID:AB_2801972 |
| IgG-PE | Miltenyi Biotec | Cat# 130-119-964  RRID:AB_2751950 |
| CD45-AF647 | Santa Cruz Biotechnology | Cat# sc-1178, RRID:AB_627074 |
| CD3-AF647 | Biolegend | Cat# 344825, RRID:AB_2563440 |
| CD163-AF647 | Biolegend | Cat# 326508, RRID:AB_893264 |
| ER-TR7-AF546 | Santa Cruz Biotechnology | Cat# sc-73355, RRID:AB_1122890 |
| Collagen I-AF555 | Bioss Antibody | Cat# bsm-33400M-A555 |
| Nucleocapsid CoV-2 | Synaptic systems | Cat.No. HS-452 011 |
| CD3-iFluor790 | AAT Bioquest, Inc | 100320M0 |
| Sytox green | Thermo Fisher | #57020 |
| **Biological samples** |  |  |
| Human autopsy lung FFPE blocks (COVID and Controls) | Department of Pathology, Charité | https://pathologie-ccm.charite.de/ |
| Human autopsy lung cryo blocks (COVID) | Department of Neuropathology | https://neuropathologie.charite.de/ |
| Human autopsy lung cryo blocks (Controls) | NeuroCure BrainBank/Biobank | https://neuropathologie.charite.de/en/research/brainbank/ |
| Human autopsy lung FFPE blocks (Controls) | NeuroCure BrainBank/Biobank | https://neuropathologie.charite.de/en/research/brainbank/ |
| Human autopsy lung draining lymph nodes cryo blocks (COVID) | Department of Neuropathology | https://neuropathologie.charite.de/ |
| Human autopsy lung draining lymph nodes cryo blocks (Controls) | NeuroCure BrainBank/Biobank | https://neuropathologie.charite.de/en/research/brainbank/ |
| **Chemicals, peptides, and recombinant proteins** | | |
| 3-aminopropyltriethoxysilane (APES) | Sigma Aldrich | CAS: 919-30-2 |
| electron microscopy grade 2% paraformaldehyde | Electron Microscopy Sciences | Cat.No. 50-980-493 |
| Quadrol solution (N,N,N′,N′-Tetrakis-(2-hydroxypropyl)-ethylendiamin) | Sigma-Aldrich | 122262 |
| CHAPS solution ((3-[3-cholamidopropyl)dimethylammonio]-1-1propanesulfonate | Sigma-Aldrich | 226947 |
| SmartClear Pro | LifeCanvas Technologies |  |
| EasyIndex | Cairn Research | DLC/EI-Z1001 |
| silica matching liquid | Cargille Laboratories | Cat#19569 |
| 2-Methylbutane 99 +% (GC) | Sigma-Aldrich Chemie AG | M32631 |
| Pikrin-Fuchsin van Gieson | Th. Geyer Berlin GmbH | 2E 050 |
| Blueing reagent | Roche | 05266769001 / 760-2037 |
| iView DAB,Benchm.Detection Kit | Roche | 05266157001 / 760-091 |
| Deparaffination solution (10x) | Roche | 05279771001 / 950-102 |
| reaction buffer (APK) (10X) | Roche | 05353955001 / 950-300 |
| Haemalaunsolution Mayer |  | ME 9249 |
| Glutaraldehyde 25% | SERVA | SVA 23115.01 |
| iron-Haematoxylin Weig A | Th. Geyer Berlin GmbH | 2E 32 |
| iron-Haematoxylin Weig B | Th. Geyer Berlin GmbH | 2E 52 |
| SHIELD 500ml kit:  SHIELD Epoxy (SH-Ex)  SHIELD Buffer (SH-BS)  SHIELD On (SH-ON) | Cairn Research | DLC/SH-500 |
| **Commercial assays** | | |
| MagNAPure 96 DNA and Viral NA Large Volume kit | Roche | 06374891001 |
| Rhonda PCR rapid COVID-19 test | Spindiag | SD003-02-020-A01 |
| Qubit dsDNA HS Assay kit | Thermo Fisher Scientific | Q32854 |
| 10x Visium Spatial Gene Expression Kit | 10x Genomics | 1000187 |
| 10x Genomics Visium Spatial Tissue Optimization Kit | 10x Genomics | 1000193 |
| Chromium Single Cell 3ʹ V3.1 library preparation kit | 10x Genomics | PN-1000121 |
| **Software and algorithms** | | |
| Atlas 5 software | Zeiss | zeiss.com/atlas5 |
| MelTec TIC-Control | MelTec GmbH & Co.KG Magdeburg, Germany |  |
| ImageJ/Fiji | Schindelin et al. 2012 | <https://doi.org/10.1038/nmeth.2019> |
| Ilastik 1.3.2 | Berg et al. 2019 | <https://doi.org/10.1038/s41592-019-0582-9> |
| CellProfiler 3.1.8 | Carpenter et al. 2006 | DOI: [10.1186/gb-2006-7-10-r100](https://doi.org/10.1186/gb-2006-7-10-r100) |
| GraphPad Prism 9.2.0 | Graph Pad Software | www.graphpad.com |
| R (version 4.1.0) | R Core Team (2020) | https://www.R-project.org/ |
| Seurat (version 4.0.4) | <https://CRAN.R-project.org/package=Seurat> | doi: [10.1038/nbt.3192](https://doi.org/10.1038/nbt.3192) |
| R (version 3.6.1) | R Core Team (2020) | https://www.R-project.org/ |
| Seurat (version 3.1.1) | <https://CRAN.R-project.org/package=Seurat> | doi: [10.1038/nbt.3192](https://doi.org/10.1038/nbt.3192) |
| Loupe Browser 5.1.0 software | 10x Genomics | https://support.10xgenomics.com/single-cell-gene-expression/software/downloads/latest |
| Space Ranger software 1.3.0 | 10x genomics | https://support.10xgenomics.com/spatial-gene-expression/software/pipelines/latest/installation |
| Gene Set Enrichment Analysis (GSEA) *fgsea* package (version 1.18.0) | Molecular Signatures Database v7.4 | [Subramanian, Tamayo, et al. (2005, PNAS)](https://www.pnas.org/content/102/43/15545) and [Mootha, Lindgren, et al. (2003, Nature Genetics)](http://www.nature.com/ng/journal/v34/n3/abs/ng1180.html). |
| single sample gene set enrichment analysis (ssGSEA) with the *escape* package (version 1.2.0) | Molecular Signatures Database v7.4 | [Subramanian, Tamayo, et al. (2005, PNAS)](https://www.pnas.org/content/102/43/15545) and [Mootha, Lindgren, et al. (2003, Nature Genetics)](http://www.nature.com/ng/journal/v34/n3/abs/ng1180.html). |
| MSigDB (v7.4) | Molecular Signatures Database v7.4 | [Subramanian, Tamayo, et al. (2005, PNAS)](https://www.pnas.org/content/102/43/15545) and [Mootha, Lindgren, et al. (2003, Nature Genetics)](http://www.nature.com/ng/journal/v34/n3/abs/ng1180.html). |
| Cell Ranger Software Suite (Version 3.1.0) | 10x genomics | https://support.10xgenomics.com/single-cell-gene-expression/software/pipelines/latest/installation |
| ImSpector software Version 7.0.73 | LaVision Biotec | LaVision BioTec GmbH  https://www.lavisionbiotec.com/ |
| Imaris Bitplanex64 Version 9.7.2. | Andor Technology Ltd. | Andor Technology Ltd.  https://imaris.oxinst.com/ |
| Imaris Stitcher Version 9.7.2. | Andor Technology Ltd. | Andor Technology Ltd.  https://imaris.oxinst.com/ |
| Imaris Converter Version 9.7.2. | Andor Technology Ltd. | Andor Technology Ltd.  https://imaris.oxinst.com/ |
| **Other** | | |
| Toponome Image Cycler® MM3 | MelTec GmbH & Co.KG Magdeburg, Germany |  |
| Superfrost Plus Gold slides | Fisher Scientific, Jena, Germany | 11976299 |
| Zeiss LSM 880 | Zeiss |  |
| MH560 cryotome | ThermoFisher |  |
| cover slides 24 x 60 mm | Menzel-Gläser | 10672-015 |
| “press-to-seal” silicone sheets | Life technologies | P24745 |
| Illumina NextSeq 500/550 | Illumina |  |
| Nextseq 500 | Illumina |  |
| Novaseq 6000 S1 | Illumina |  |
| Makro fluoreszenz-cuvette | Msscientific Chromatographie-Handel GmbH, Berlin, Germany | FF-1FL-G-5 |
| Ultramicroscope UM-0108 | LaVision BioTec |  |
| LifeCanvas SmartClear II Pro | Cairn Research,  Faversham, United Kingdom | https://lifecanvastech.com/products/smartclear-ii-pro/ |
| Cork plates, 20mm | Slee medical GmbH | #30001001 |
| Glas slides, 76 x 26, SuperFrost Plus | Langenbrink | #041300 |
| Embedding media Paraplast Plus | Roth | X881.1 |
| OCT Compound, Tissue Tek | Charité | AK 4583 SS |
